# Associations between prenatal adversity and neonatal white matter microstructure on language outcomes at age 2 years

**DOI:** 10.1101/2024.08.02.24311434

**Authors:** Jacob Bjork, Jeanette K. Kenley, Caleb Gardner, Aidan Latham, Tara A. Smyser, J. Philip Miller, Joshua J. Shimony, Jeffrey J. Neil, Barbara Warner, Joan Luby, Deanna M. Barch, Cynthia E. Rogers, Christopher D. Smyser, Rachel E. Lean

## Abstract

**Background:** Early life adversity is associated with microstructural alterations in white matter regions that subserve language. However, the mediating and moderating pathways between adversities experienced *in utero* and key neonatal white matter tracts including the corpus callosum (CC), superior longitudinal fasciculus (SLF), arcuate fasciculus (AF), inferior fronto-occipital fasciculus (IFOF), and uncinate on early language outcomes remains unknown.

**Methods:** This longitudinal study includes 160 neonates, oversampled for prenatal exposure to adversity, who underwent diffusion MRI (dMRI) in the first weeks of life. dMRI parameters were obtained using probabilistic tractography in FSL. Maternal Social Disadvantage and Psychosocial Stress was assessed throughout pregnancy. At age 2 years, the Bayley Scales of Infant and Toddler Development-III evaluated language outcomes. Linear regression, mediation, and moderation assessed associations between prenatal adversities and neonatal white matter on language outcomes.

**Results:** Prenatal exposure to Social Disadvantage (p<.001) and Maternal Psychosocial Stress (p<.001) were correlated with poorer language outcomes. When Social Disadvantage and maternal Psychosocial Stress were modeled simultaneously in relation to language outcomes, only Social Disadvantage was significant (p<.001). Independent of Social Disadvantage (p<.001), lower neonatal CC fractional anisotropy (FA) was related to poorer global (p=.02) and receptive (p=.02) language outcomes. CC FA did not mediate the association between Social Disadvantage and language outcomes (indirect effect 95% CIs -0.96-0.15), and there was no interaction between Social Disadvantage and CC FA on language outcomes (p>.05). Bilateral SLF/AF, IFOF, and uncinate were not significant (p>.05).

**Conclusions:** Prenatal exposure to Social Disadvantage and neonatal CC FA were independently related to language problems by age 2, with no evidence of mediating or moderating associations with language outcomes. These findings elucidate the early neural underpinnings of language development and suggest that the prenatal period may be an important time to provide poverty- reducing support to expectant mothers to promote offspring neurodevelopmental outcomes.

---

Speech and language delays are one of the most common neurodevelopmental problems, affecting 8 – 20% of school-age children (Nelson, Nygren, Walker, & Panoscha, 2006). Early childhood is a critical period of language development. Receptive language skills, such as the discrimination of phonemes, begin to emerge in the first year of life (Gervain, 2020). Expressive language skill acquisition also rapidly unfolds during early childhood, with children progressing from basic two-word sentences in older infancy to storytelling and compound sentences by age 4 years (Visser-Bochane, Reijneveld, Krijnen, van der Schans, & Luinge, 2020). Importantly, receptive and expressive language deficits pose significant challenges for children across multiple domains, including school readiness delays, communication-interaction problems, and socioemotional dysfunction (Bornstein et al., 2020; Chow, Ekholm, & Coleman, 2018; Kastner, May, & Hildman, 2001; Vallotton & Ayoub, 2011). Investigation of the mechanisms that underlie disparities in language development is important to identify children in need of early language interventions and improve longer-term literacy and social-communication outcomes.

Childhood exposure to social disadvantage and maternal psychosocial stress are well-established, and relatively stable, risk factors for language delays throughout childhood (Fernald, Marchman, & Weisleder, 2013; Justice et al., 2019; Perkins, Finegood, & Swain, 2013). While such postnatal exposures play a critical role in disrupting language skill acquisition, exposures during the prenatal period may also be important (Gervain, 2020). In a retrospective cohort study of infants born preterm, children born to mothers residing in disadvantaged neighborhoods during pregnancy showed higher rates of language delays and impairments in early childhood (Nwanne et al., 2022). Similarly, aspects of maternal psychosocial dysfunction including depression, anxiety, and stress during pregnancy are also related to poorer language outcomes in childhood (D’Souza et al., 2019; Laplante, Brunet, Schmitz, Ciampi, & King, 2008; Naudé et al., 2022). However, these previous investigations did not oversample for exposure to adversity, employed a retrospective study design, and/or assessed language outcomes in infants born preterm. Given these considerations, associations between prenatal adversity and early language development remain unclear.

Prenatal programming posits that adversity experienced *in utero* has a pronounced impact on vulnerable fetal structural and functional brain systems and, in turn, increases offspring susceptibility for adverse neurodevelopmental outcomes (Kwon & Kim, 2017). The prenatal environment has been linked with variability in newborn neural responses to speech heard in the womb, suggesting that prenatal experiences begin to shape language circuitry *in utero* when axonal development and myelination may be especially vulnerable to intra-uterine experiences (Dubois et al., 2014; Gervain, 2015). Prenatal exposure to social disadvantage and maternal psychosocial stress has been associated with lower fractional anisotropy (FA) in the corpus callosum (CC) and uncinate in neonates (Graham et al., 2020; Lautarescu, Craig, & Glover, 2020; Thompson et al., 2019). Longitudinal follow-up studies also suggest that neonatal microstructural alterations in the CC, uncinate, inferior fronto-occipital fasciculus (IFOF), and longitudinal fasciculi increase the likelihood of language problems as early as age 12 months (Bugada, Kline, & Parikh, 2021; Dubner, Rose, Bruckert, Feldman, & Travis, 2020; Feng et al., 2019; Salvan et al., 2017; Sket et al., 2019), with similar cross-sectional findings in school-age children (Mürner-Lavanchy et al., 2018; Ostertag et al., 2023). However, no study has yet examined the extent that neonatal alterations in language pathways represent a key neural mechanism by which prenatal adversity places children at risk of language delays in early childhood.

In the Early Life Adversity and Biological Embedding (eLABE) cohort, we have previously shown that prenatal social disadvantage (PSD) is related to altered neonatal CC, uncinate, and IFOF microstructure (Lean et al., 2022; Sanders et al., 2024). Research from other groups also demonstrates prospective associations between disrupted neonatal white matter organization and difficulties in early language skill acquisition (Dubner et al., 2020; Feng et al., 2019; Sket et al., 2019). Taken together, these previous findings might suggest that adversity-related alterations in neonatal white matter may be a key mediator linking prenatal adversity with language delays in the first years of life. Prenatal exposure to adversity may also moderate associations between white matter organization and language development. In the eLABE cohort, associations between prenatal exposure to measures of maternal perceived or biological stress were differentially related to neonatal macro- or micro-structural brain outcomes depending on the level of PSD, with disrupted or maladaptive relationships among dyads experiencing higher levels of PSD (Herzberg et al., 2023; Lean et al., 2022; Sanders et al., 2024). However, the extent that prenatal adversity disrupts or exacerbates links between altered neonatal white matter and early language problems remains unknown.

This study seeks to address this gap in by examining prenatal exposure to adversity and neonatal white matter microstructure in relation to language outcomes at age 2 years in a cohort recruited prenatally and oversampled for adversity. Current study aims were to examine associations between maternal social and psychosocial adversity during pregnancy on child language outcomes at age 2 years, identify the neonatal white matter correlates of early language outcomes focusing on the CC, IFOF, uncinate, and a bundle comprised of the superior longitudinal fasciculus/arcuate fasciculus (SLF/AF, see Supporting Information) as key language pathways, and investigate the mediating and moderating associations between prenatal adversity, neonatal white matter microstructure, and language outcomes. We hypothesized that neonatal white matter microstructure would mediate the link between prenatal adversity and language outcomes and that associations between neonatal white matter and language outcomes would be disrupted in children experiencing higher levels of adversity (Lean et al., 2022; Luby et al., 2024; Sanders et al., 2024).

## METHODS

### Sample & Ethical Considerations

The current study draws data from the longitudinal eLABE cohort of 398 mother-child dyads (Luby et al., 2023). From 09/2017 to 02/2020, 395 women were recruited during pregnancy (four mothers had two singleton pregnancies) identified from the Washington University March of Dimes Prematurity Research Center (Stout et al., 2022). Mothers were oversampled from an obstetric clinic serving low-income women to enrich the cohort for adversity. eLABE exclusion criteria were multiple gestation pregnancies, congenital infections, and prenatal alcohol or substance use (other than tobacco or marijuana for representativeness of pregnant women experiencing adversity (el Marroun et al., 2008; Passey, Sanson-Fisher, D’Este, & Stirling, 2014)). Study procedures were approved by the Washington University Institutional Review Board. Written informed consent was obtained from all mothers.

### Procedure

Mothers completed surveys in each trimester of pregnancy to assess social background and mental health. At an average of 41 weeks postmenstrual age (PMA, range 38-45 weeks), non-sedated neonates underwent multimodal MRI scans on a Siemens Prisma 3T Scanner with a 64- channel head coil. Neonates were fed, swaddled, and positioned in a head-stabilized vacuum wrap before being placed in the head coil on foam padding to reduce motion. At age 2 years, 206 children returned for a neurodevelopmental assessment which included the Bayley Scales of Infant and Toddler Development-III (Bayley-III). Paired neonatal diffusion magnetic resonance imaging (dMRI) and 2-year language outcome data were obtained for 160 children (Table 1, Figure S1). Infants with high-grade perinatal brain injury and/or admission to the neonatal intensive care unit >7 days were excluded from the current study analyses.

**Table 1.**
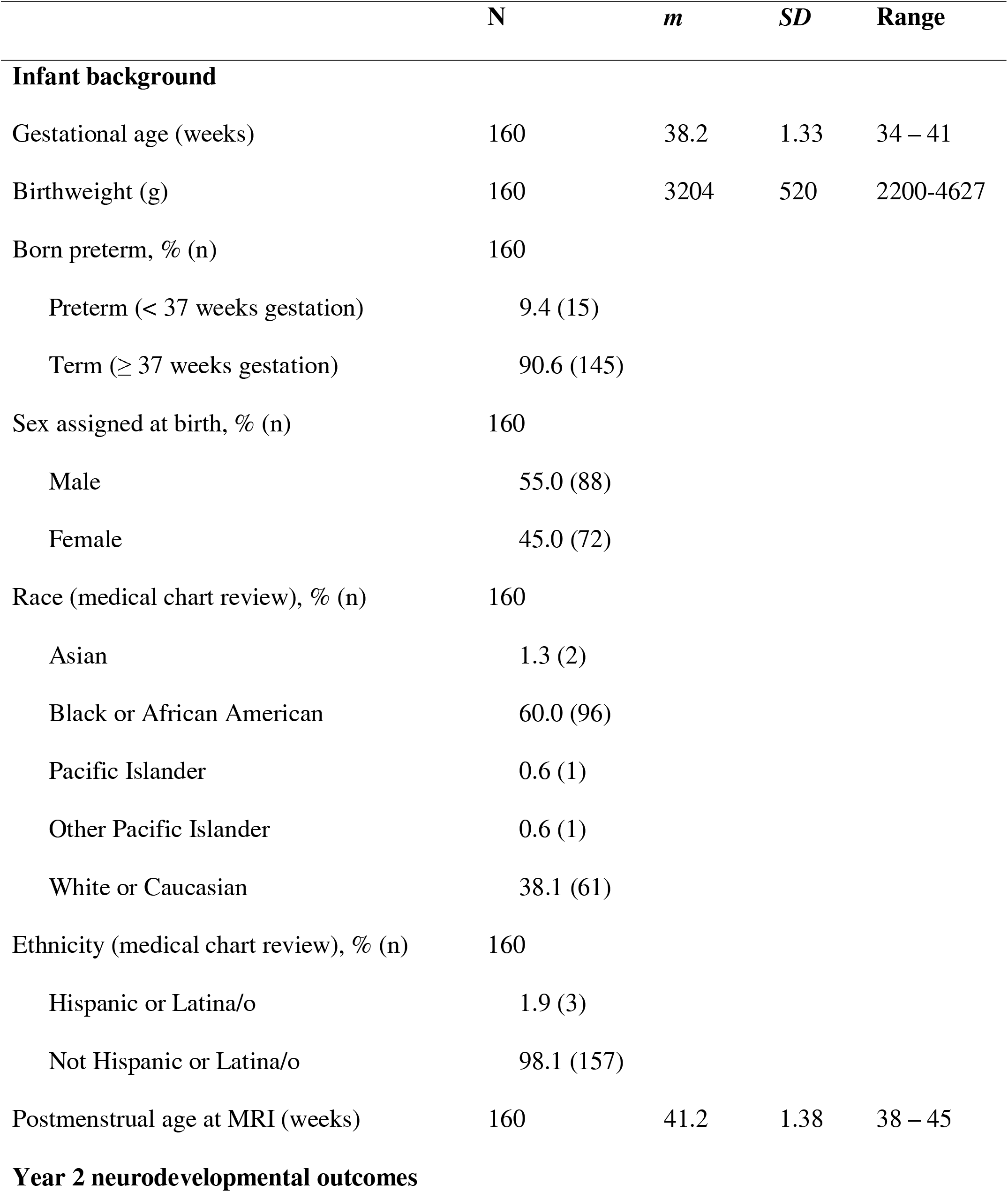
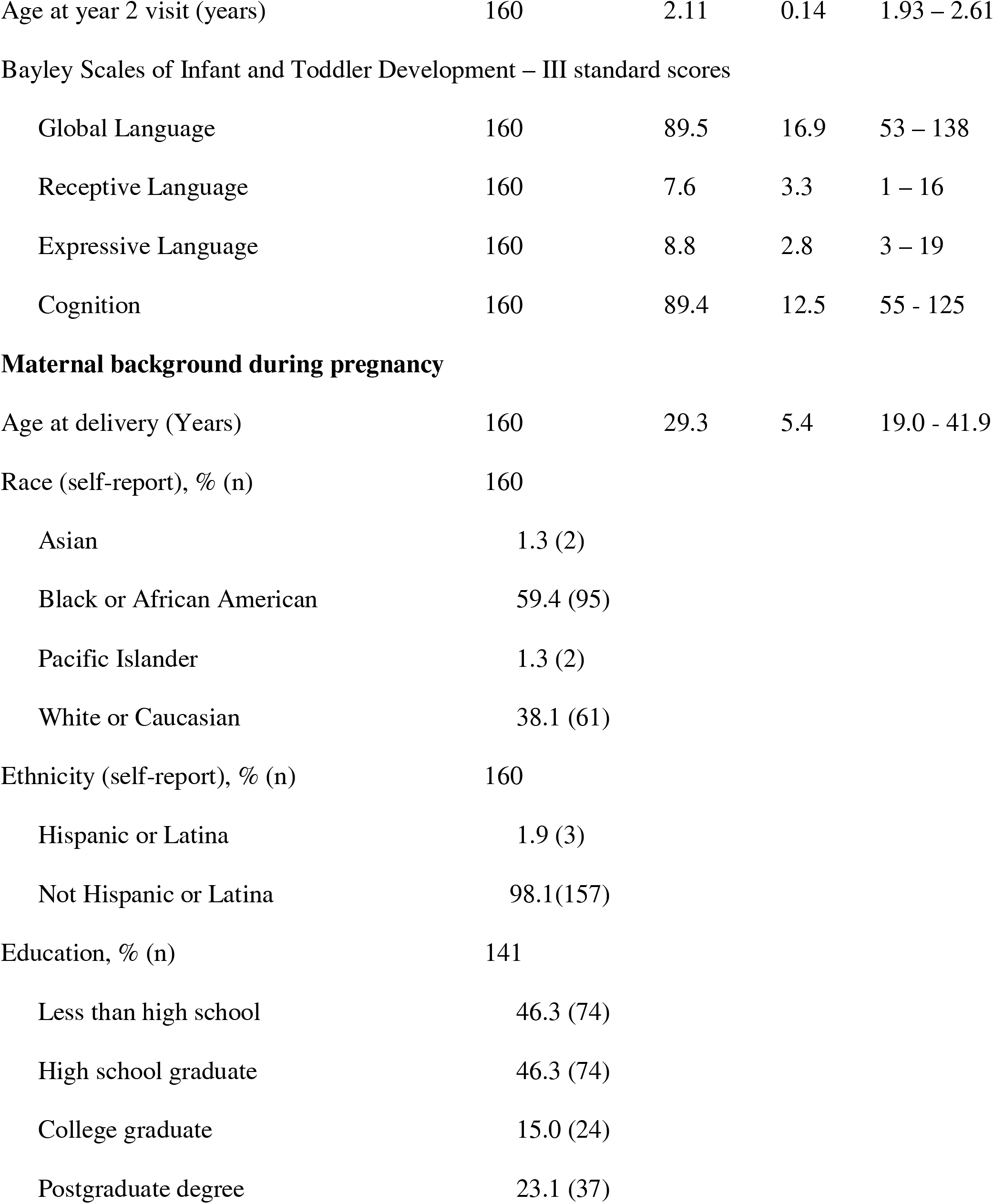
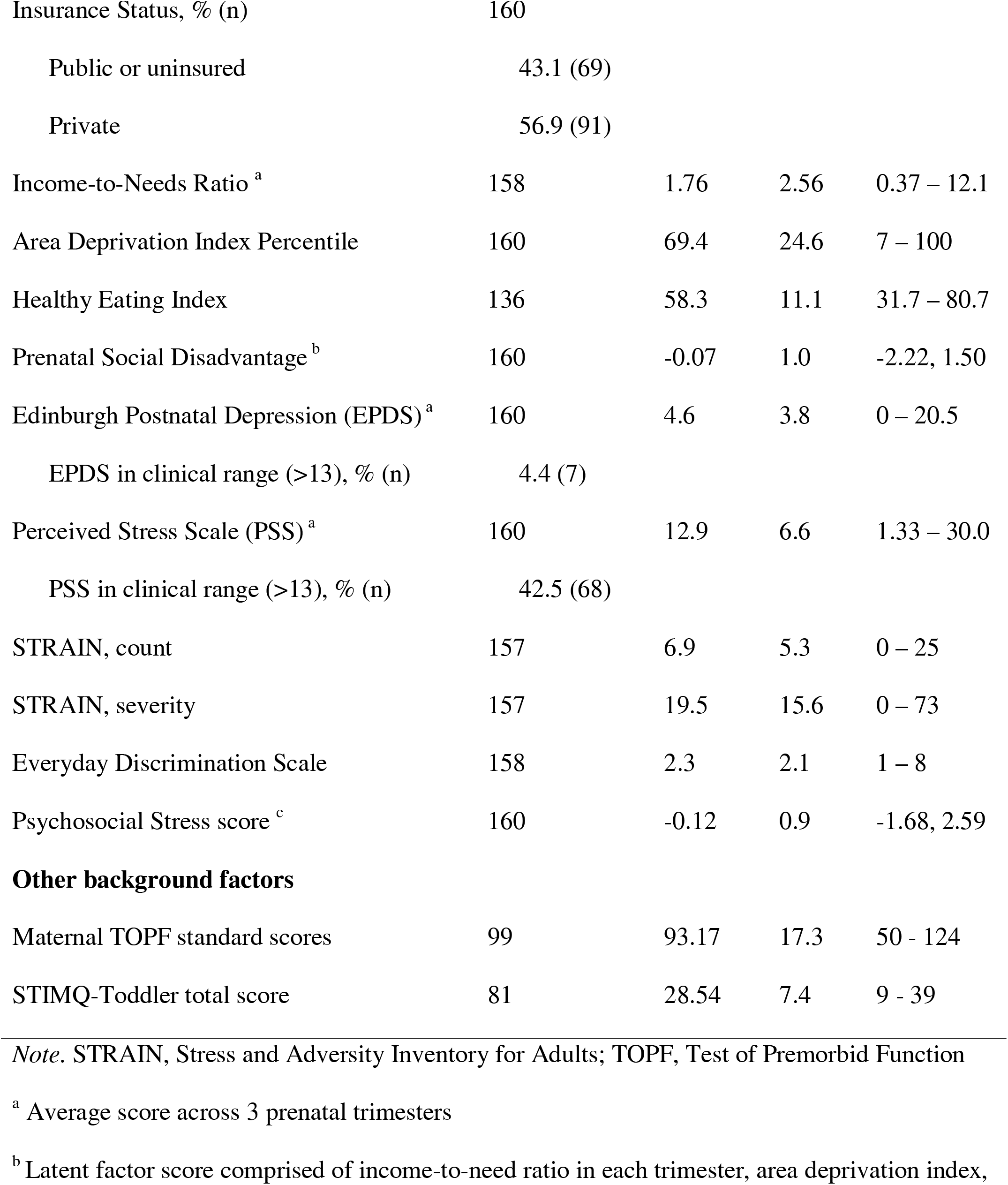
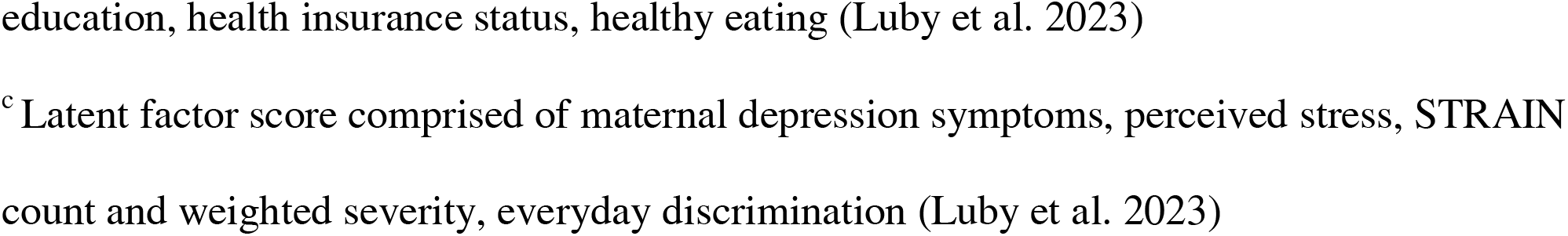
Sample background characteristics (*n*=160)

### Measures

#### Prenatal Adversities

As described in Luby et al. (2023), structural equation modeling was used in the larger eLABE cohort to derive two latent constructs of prenatal adversity. First, PSD provided an aggregated measure of financial and material hardship (Luby et al., 2023). Measures included maternal health insurance status, highest level of education, family income-to-needs ratios (Office of the Assistant Secretary for Planning and Evaluation, 2024), Area Deprivation Index (Kind & Buckingham, 2018), and the Healthy Eating Index-2015 (Krebs-Smith et al., 2018). Observed social variables were highly correlated (-0.66-0.74, p<.001), supporting the use of a latent factor score.

Second, Psychosocial Stress was comprised of maternal depression symptoms, perceived stress, stressful/traumatic events, and racial discrimination (Luby et al., 2023). Measures included the Edinburgh Postnatal Depression Scale (Cox, Holden, & Sagovsky, 1987), Cohen Perceived Stress Scale (Cohen, Kamarck, & Mermelstein, 1983), Stress and Adversity Inventory for Adults (Slavich & Shields, 2018), and Everyday Discrimination Survey (Williams, Yu, Jackson, & Anderson, 1997). All psychosocial stress variables were correlated (0.21-0.93, p<0.01) (Luby et al., 2023). In the current study sample, PSD and Psychosocial Stress were moderately correlated (r=0.40, p<.001). Similar Social Disadvantage and Psychosocial Stress factors were also created for the 2-year follow-up (Table S1).

#### Neonatal White Matter Tracts

Neonatal dMRI scans were acquired as two 5-minute runs using MB4, TR/TE=2500/79.4 ms, (1.75 mm)^3^ voxels, with whole brain coverage (80 slices), 108 b-values sampled on three shells of b=500-2500 s/mm^2^ and 7 b=0 images interspersed throughout each run with phase encoding direction reversal (anterior→posterior and posterior→anterior) for susceptibility- and eddy-current distortion correction. Seed regions-of-interest (ROIs) were created for the start-, way-, and end-points of each white matter tract using standard anatomical landmarks on a neonatal FA atlas. The atlas was then registered to the subject’s native space FA image using FSL FLIRT (version 6.0.4). Calculated transforms were applied to each set of seed ROIs. Probabilistic tractography was performed using FSL Probtrackx. The diffusion tensor model was completed using FSL’s dtifit. Tensors were fitted using FSL’s bedpostx and modelled to allow for two crossing fibers. If more than one waypoint mask was required, waypoint crossing was forced in listed order. Probtrackx output files were then thresholded to retain streamlines indicating greater certainty of white matter. All tracts were visually and numerically inspected. Infants whose data were consistently outside of 3 standard deviations (SD) were adjusted as required. Key tracts of interest (Figure 1) included the CC, IFOF, uncinate, and a SLF/AF language bundle. The SLF and the superior portion of the AF were examined as a language bundle given the proximity of the SLF and AF (Figures S2-S3 demonstrating tract overlap) and the relative immaturity of these tracts in neonates which precluded the isolation of the SLF and AF as separate pathways (Liang et al., 2022; Schilling et al., 2022; Zhang et al., 2006). The corticospinal tract (CST) was included as a negative control tract (Natali, Reddy, & Bordoni, 2024).

**Figure 1.**
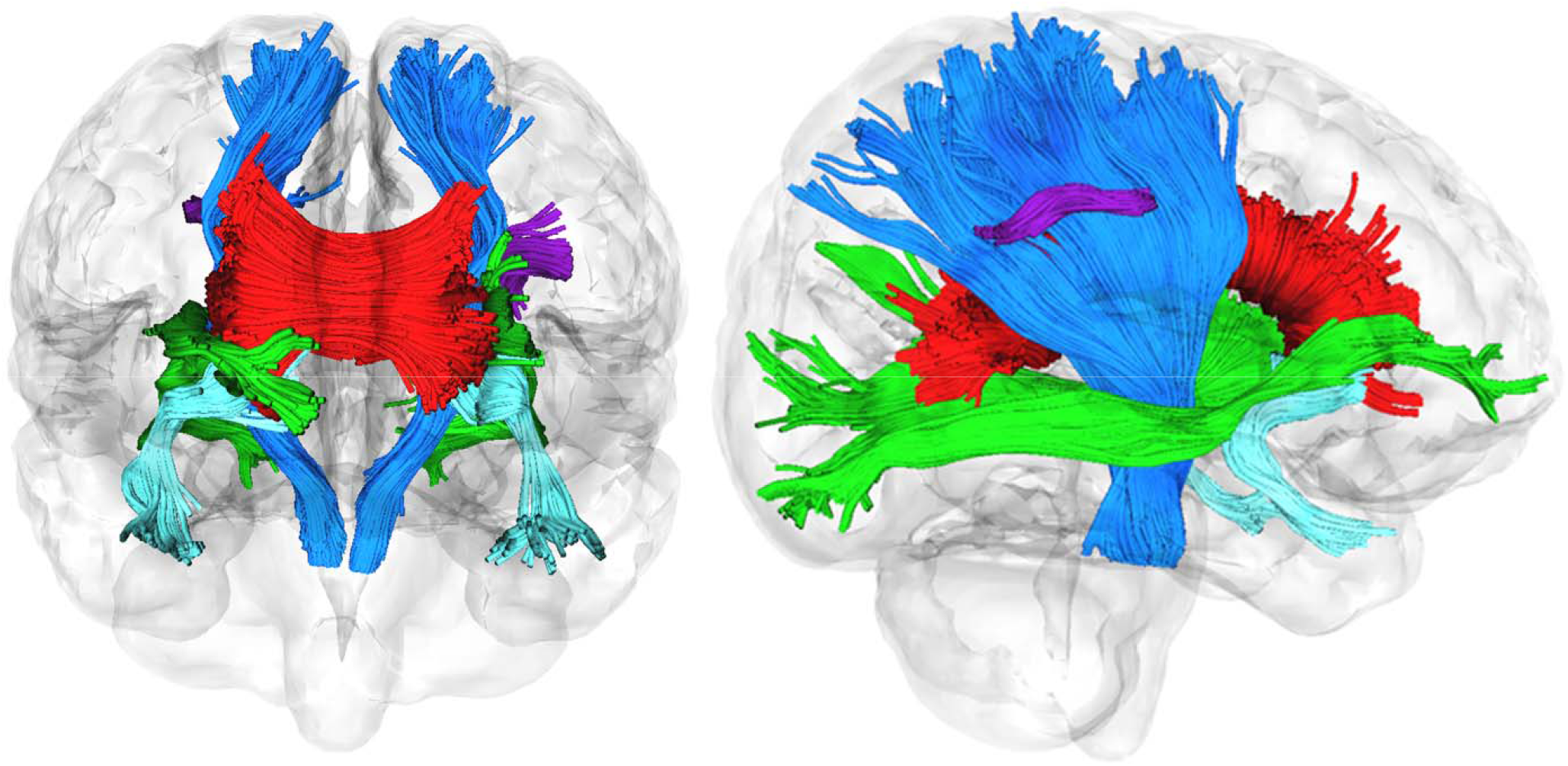
Illustration of key white matter tracts of interest. Key tracts of interest involved in language pathways include the uncinate (aqua), corpus callosum (red), inferior occipital fasciculus (green), and superior longitudinal fasciculus/arcuate fasciculus/language bundle (purple). The corticospinal tract (blue) was included as a negative control tract.

#### Language at Age 2 Years

The Bayley-III was administered by psychometricians at the 2-year follow-up to evaluate children’s language and cognitive development. Age-normed standard scores for global language and scaled scores for receptive and expressive language are reported. As some participants completed follow-up before the COVID-19 Pandemic Lockdown (n=29, 18%), between-groups differences in timing of language assessment before and after March 16^th^, 2020 were conducted, with no differences in outcomes found (Table S2).

#### Maternal Cognitive Ability and Home Enrichment

Maternal cognitive ability was assessed using the Test of Premorbid Function (TOPF) (Wechsler, 2012). Age-normed standard scores are reported (data collected n=99, 62%). The TOPF demonstrates concurrent validity with the Wechsler Adult Intelligence Scale-IV (Wechsler, 2012). The parent-report StimQ-Toddler was also completed at the 2-year follow-up as a measure of cognitive enrichment provided in the home, assessing availability of learning materials, reading books, parental involvement in developmental advancement, and parental verbal responsiveness (Cates et al., 2023). StimQ-Toddler total scores are reported (data collected n=81, 51%). There was no difference in PSD between participants who did or did not have TOPF (*t*=0.49, *p*=.62) or StimQ-Toddler (*t*=-0.50, *p*=.62) data collected.

### Data Analysis

Data analysis was performed in SPSS (Version 28). Neonatal dMRI variables (FA, mean diffusivity [MD], axial diffusivity [AD], and radial diffusivity [RD]) and child language outcome scores (global, receptive, and expressive) were screened for non-normal distributions and extreme outlier values (>3 SD). Extreme outliers were corrected by assigning the next poorest non-extreme outlier score in the distribution.

Linear regression models were used to relate PSD and maternal Psychosocial Stress to child language outcomes, adjusted for gestational age (GA) and sex assigned at birth (male=1, female=2). For each dependent variable (global, receptive, and expressive language scores), latent prenatal adversity constructs were first fitted separately as independent variables, and then simultaneously to evaluate the independent contribution of each latent factor. A mean-centered interaction term between PSD and Psychosocial Stress was also tested. Regression models were also performed with dMRI variables as independent variables, along with PMA at MRI scan, to identify neonatal white matter correlates of language outcomes. Regression models were BH-False Discovery Rate corrected for multiple comparisons (q<.05). The PROCESS 4.0 macro for SPSS (Hayes & Rockwood, 2017) was used to evaluated whether neonatal white matter mediated (95% confidence intervals performed with bootstrapping, 5000 samples) the association between prenatal adversity and language outcomes. Formal moderation analysis with mean-centered interaction terms examined whether associations between neonatal white matter and language outcomes were moderated by PSD.

Additional analyses were performed including TOPF standard scores and StimQ-Toddler total scores as independent variables to account for maternal cognitive ability and home enrichment, respectively. Analysis was also performed with Bayley-III Cognition standard scores as the dependent variable to examine the specificity of dMRI findings for outcomes. Last, sensitivity analysis was undertaken excluding children with significant cognitive delay (Bayley-III Cognition <70, n=17) and children born preterm (<37 weeks GA, n=15).

## RESULTS

### Prenatal Adversities and Language Outcomes

When modeled individually, PSD (p<.001; total R^2^ range .17 to .27) and Psychosocial Stress (p≤.01; total R^2^ range .07 to .14) were each associated with global, receptive, and expressive language outcomes by age 2, after accounting for sex and GA (Table 2). However, when both prenatal latent factors were simultaneously entered in the regression models, only PSD remained significant (p<.001). There was no interaction between PSD and maternal Psychosocial Stress on language outcomes (all p>.05).

**Table 2.**
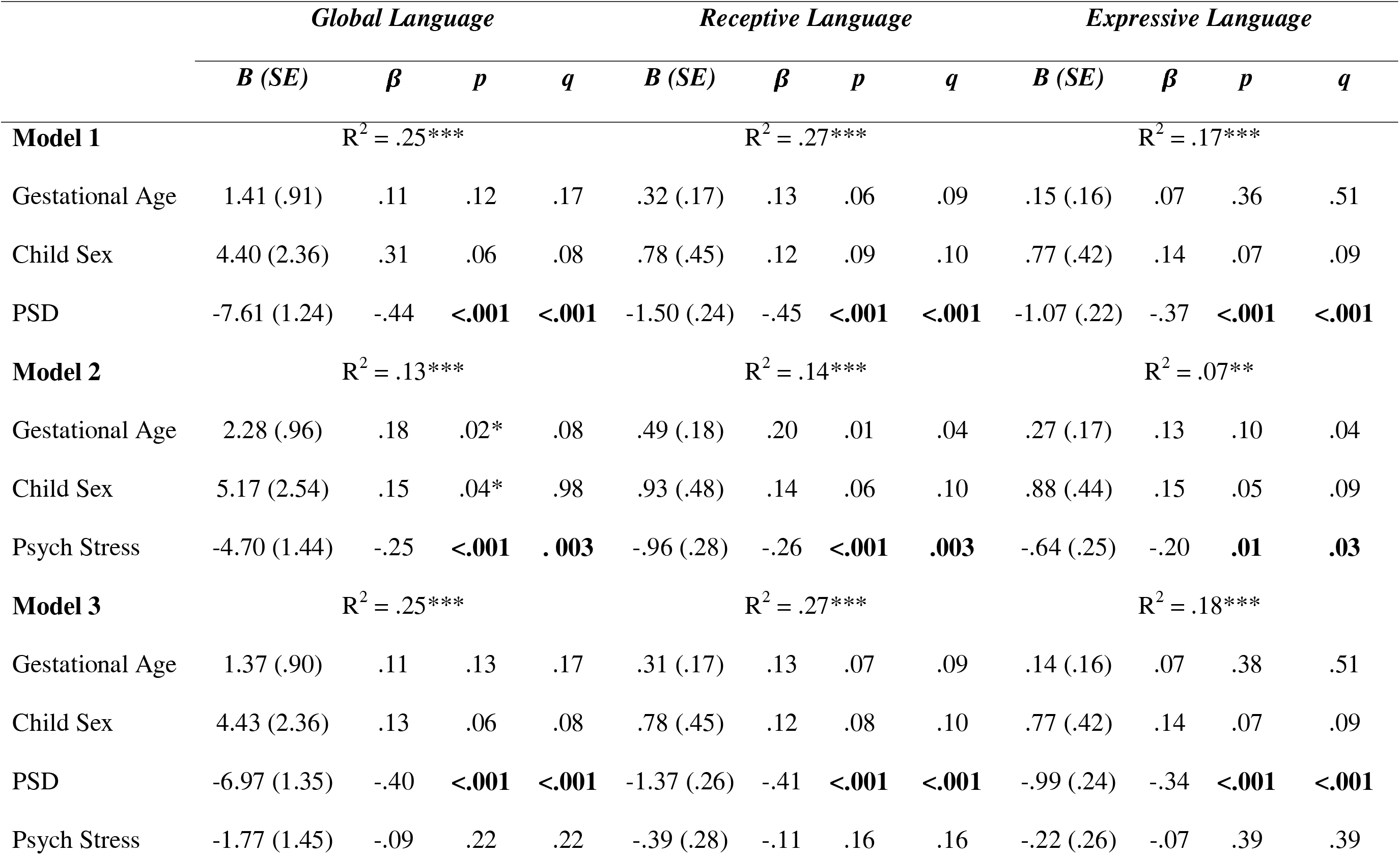
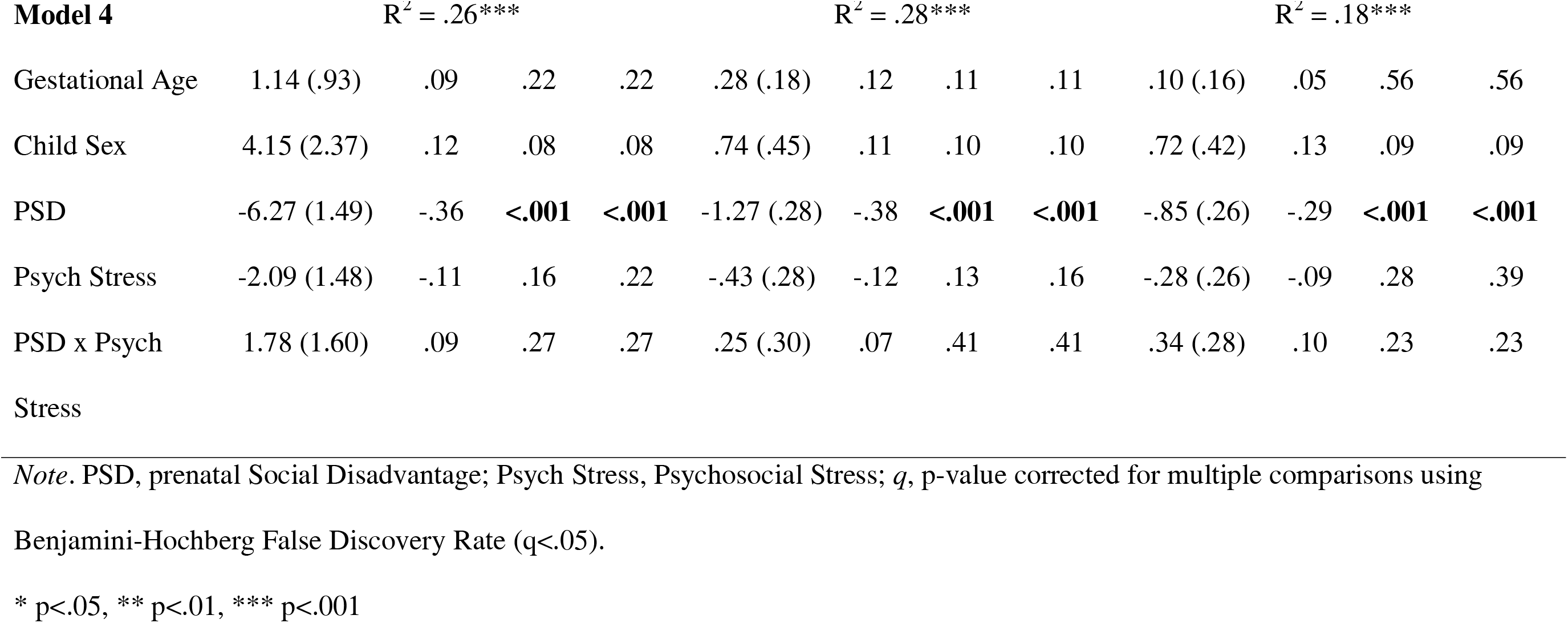
Associations between prenatal adversity and language outcomes at age 2 years

As expected, year 2 Social Disadvantage was also correlated with language ability at age 2 years (Pearson’s r range -.38 to -.44, p<.001, Table S1). However, prenatal and year 2 Social Disadvantage were highly colinear (r=.93, p<.001) and could not be included simultaneously in regression models. Given our interest in prospective associations between prenatal adversity, neonatal white matter, and language outcomes, PSD was included in subsequent regression and mediation analyses. However, moderation analyses were undertaken first with PSD, and then with 2-year Social Disadvantage, to examine whether the timing of exposure to Social Disadvantage impacted moderation analyses.

### Neonatal White Matter and Language Outcome

White matter tract descriptives are shown in Table S3. Results of regression models (Table 3) showed that lower neonatal CC FA (p=.01) and CC AD (p=.02) were associated with poorer global language ability by age 2, after covariate adjustment. Similarly, lower CC FA (p=.01) and AD (p=.02) were related to poorer receptive language outcomes (Table S4), whereas only lower CC FA (p=.02) was related to poorer expressive outcomes (Table S5). CC MD and RD were not associated with language outcomes (p>.05, Table S6). There were no significant findings for the other hypothesized tracts including the SLF/AF language bundle, IFOF, or uncinate (p>.05, Table 3). The negative control CST was also not related to language outcomes (p>.05, Table 3).

**Table 3.** Associations between neonatal white matter microstructure and global language outcomes at age 2 years.

|  | *FA* |  |  |  | *AD* |  |  |  |
| --- | --- | --- | --- | --- | --- | --- | --- | --- |
| | *B (SE)* | $\beta$ | *p* | *q* | *B (SE)* | $\beta$ | *p* | *q* |
| **Model 1: Corpus Callosum** | $R^2 = .12^{***}$ | | | | $R^2 = .10^{**}$ | | | |
| Gestational Age | 2.08 (1.03) | .16 | .05 | .05 | 1.88 (1.05) | .15 | .08 | .08 |
| Child Sex | 5.94 (2.57) | .18 | .02 | .05 | 6.08 (2.61) | .18 | .02 | .06 |
| PMA at MRI scan | -.20 (1.08) | -.02 | .85 | .85 | 1.64 (1.04) | .13 | .12 | .39 |
| Corpus Callosum | 136.9 (49.4) | .24 | **.01** | **.01** | 107.8 (47.6) | .18 | **.02** | **.02** |
| **Model 2: R SLF/AF** | $R^2 = .07^*$ | | | | $R^2 = .05^*$ | | | |
| Gestational Age | 2.30 (1.07) | .18 | .03 | .05 | 2.18 (1.06) | .17 | .04 | .06 |
| Child Sex | 5.30 (2.62) | .16 | .05 | .05 | 5.13 (2.63) | .15 | .05 | .06 |
| PMA at MRI scan | 1.02 (1.10) | .08 | .36 | .78 | .69 (1.10) | .06 | .52 | .52 |
| R SLF/AF | -3.52 (48.6) | -.01 | .94 | .93 | -7.78 (10.9) | -.06 | .48 | .48 |
| **Model 3: L SLF/AF** | $R^2 = .05^*$ | | | | $R^2 = .05^*$ | | | |
| Gestational Age | 2.26 (1.07) | .18 | .04 | .05 | 2.27 (1.06) | .18 | .03 | .06 |
| Child Sex | 5.29 (2.62) | .16 | .05 | .05 | 5.29 (2.63) | .16 | .05 | .06 |

|  | *FA* |  |  |  | *AD* |  |  |  |
| --- | --- | --- | --- | --- | --- | --- | --- | --- |
| | *B (SE)* | $\beta$ | *p* | *q* | *B (SE)* | $\beta$ | *p* | *q* |
| PMA at MRI scan | .91 (1.16) | .07 | .43 | .78 | .95 (1.12) | .08 | .40 | .51 |
| L SLF/AF | 7.20 (51.0) | .01 | .89 | .89 | -1.08 (12.5) | -.01 | .93 | .93 |
| **Model 4: R Uncinate** | $R^2 = .07^*$ | | | | $R^2 = .08^*$ | | | |
| Gestational Age | 2.29 (1.06) | .16 | .03 | .05 | 2.43 (1.07) | .19 | .02 | .06 |
| Child Sex | 5.28 (2.63) | .09 | .05 | .05 | 5.05 (2.64) | .15 | .06 | .06 |
| PMA at MRI scan | 1.06 (1.11) | -.01 | .34 | .78 | .76 (1.05) | .06 | .47 | .52 |
| R Uncinate | -8.68 (55.8) | .03 | .88 | .88 | -24.5 (30.9) | -.06 | .43 | .43 |
| **Model 5: L Uncinate** | $R^2 = .07^*$ | | | | $R^2 = .07^*$ | | | |
| Gestational Age | 2.36 (1.07) | .19 | .03 | .05 | 2.23 (1.07) | .18 | .05 | .06 |
| Child Sex | 5.24 (2.63) | .16 | .05 | .05 | 5.30 (2.62) | .16 | .05 | .06 |
| PMA at MRI scan | 1.15 (1.08) | .09 | .29 | .78 | 1.07 (1.05) | .09 | .31 | .51 |
| L Uncinate | -24.8 (55.9) | -.04 | .66 | .66 | 81.2 (41.9) | .16 | .05 | .05 |
| **Model 6: R IFOF** | $R^2 = .07^*$ | | | | $R^2 = .09^{**}$ | | | |

|  | *FA* |  |  |  | *AD* |  |  |  |
| --- | --- | --- | --- | --- | --- | --- | --- | --- |
| | *B (SE)* | $\beta$ | *p* | *q* | *B (SE)* | $\beta$ | *p* | *q* |
| Gestational Age | 2.24 (1.06) | .18 | .04 | .05 | 2.12 (1.05) | .17 | .04 | .06 |
| Child Sex | 5.30 (2.62) | .16 | .05 | .05 | 5.97 (2.62) | .18 | .02 | .06 |
| PMA at MRI scan | .73 (1.24) | .06 | .56 | .84 | 1.82 (1.09) | .15 | .10 | .39 |
| R IFOF | 17.8 (48.1) | .04 | .71 | .71 | 81.2 (41.9) | .16 | .05 | .05 |
| **Model 7: L IFOF** | | $R^2 = .07^*$ | | | | $R^2 = .09^{**}$ | | |
| Gestational Age | 2.29 (1.06) | .18 | .03 | .05 | 2.09 (1.06) | .17 | .05 | .06 |
| Child Sex | 5.27 (2.64) | .16 | .05 | .05 | 5.96 (2.65) | .18 | .03 | .06 |
| PMA at MRI scan | 1.04 (1.22) | .09 | .39 | .78 | 1.70 (1.12) | .14 | .13 | .39 |
| L IFOF | -3.96 (49.0) | -.01 | .94 | .94 | 62.1 (42.0) | .13 | .14 | .14 |
| **Model 8: R CST** | | $R^2 = .08^*$ | | | | $R^2 = .07^*$ | | |
| Gestational Age | 2.10 (1.07) | .17 | .05 | .05 | 2.30 (1.07) | .18 | .03 | .06 |
| Child Sex | 5.36 (2.62) | .16 | .04 | .05 | 5.30 (2.63) | .16 | .05 | .06 |
| PMA at MRI scan | .51 (1.13) | .04 | .65 | .84 | 1.04 (1.13) | .09 | .36 | .51 |

|  | *FA* |  |  |  | *AD* |  |  |  |
| --- | --- | --- | --- | --- | --- | --- | --- | --- |
| | *B (SE)* | $\beta$ | *p* | *q* | *B (SE)* | $\beta$ | *p* | *q* |
| R CST | 80.4 (84.0) | .09 | .34 | .34 | 2.11 (18.7) | .01 | .91 | .91 |
| **Model 9: L CST** | | $R^2 = .08^*$ | | | | $R^2 = .07^*$ | | |
| Gestational Age | 2.08 (1.06) | .16 | .05 | .05 | 2.34 (1.07) | .18 | .03 | .06 |
| Child Sex | 5.52 (2.62) | .16 | .04 | .05 | 5.34 (2.63) | .16 | .05 | .06 |
| PMA at MRI scan | .37 (1.14) | .03 | .75 | .84 | 1.16 (1.14) | .10 | .31 | .51 |
| L CST | 93.6 (78.3) | .11 | .23 | .23 | 6.40 (19.1) | .03 | .74 | .74 |

*Note.* PSD, prenatal Social Disadvantage; PMA, post-menstrual age; R, right; L, left; SLF/AF, superior longitudinal fasciculus/arcuate fasciculus; IFOF, inferior fronto-occipital fasciculus; CST, corticospinal tract; *q*, p-value corrected for multiple comparisons using Benjamini-Hochberg False Discovery Rate ( $q < .05$ ).
\* $p < .05$ , \*\* $p < .01$ , \*\*\* $p < .001$

### PSD and Neonatal White Matter Microstructure on Language Outcomes

Results of stepwise linear regression analysis showed that PSD (p<.001, Step 1 R^2^=.25) and lower neonatal CC FA (p=.02, Step 2 R^2^ change=.03) were independently associated with poorer global language outcomes at age 2 years (Table 4). Similar results for PSD (p<.001, Step 1 R^2^=.27) and CC FA (p=.02, Step 2 R^2^ change=.03) were observed for receptive language skills, whereas only PSD (p<.001, Step 1 R^2^=.17) was associated with expressive language skills. CC AD was not associated with language outcomes (p>.05) after accounting for PSD (p<.001, Table S7).

**Table 4.**
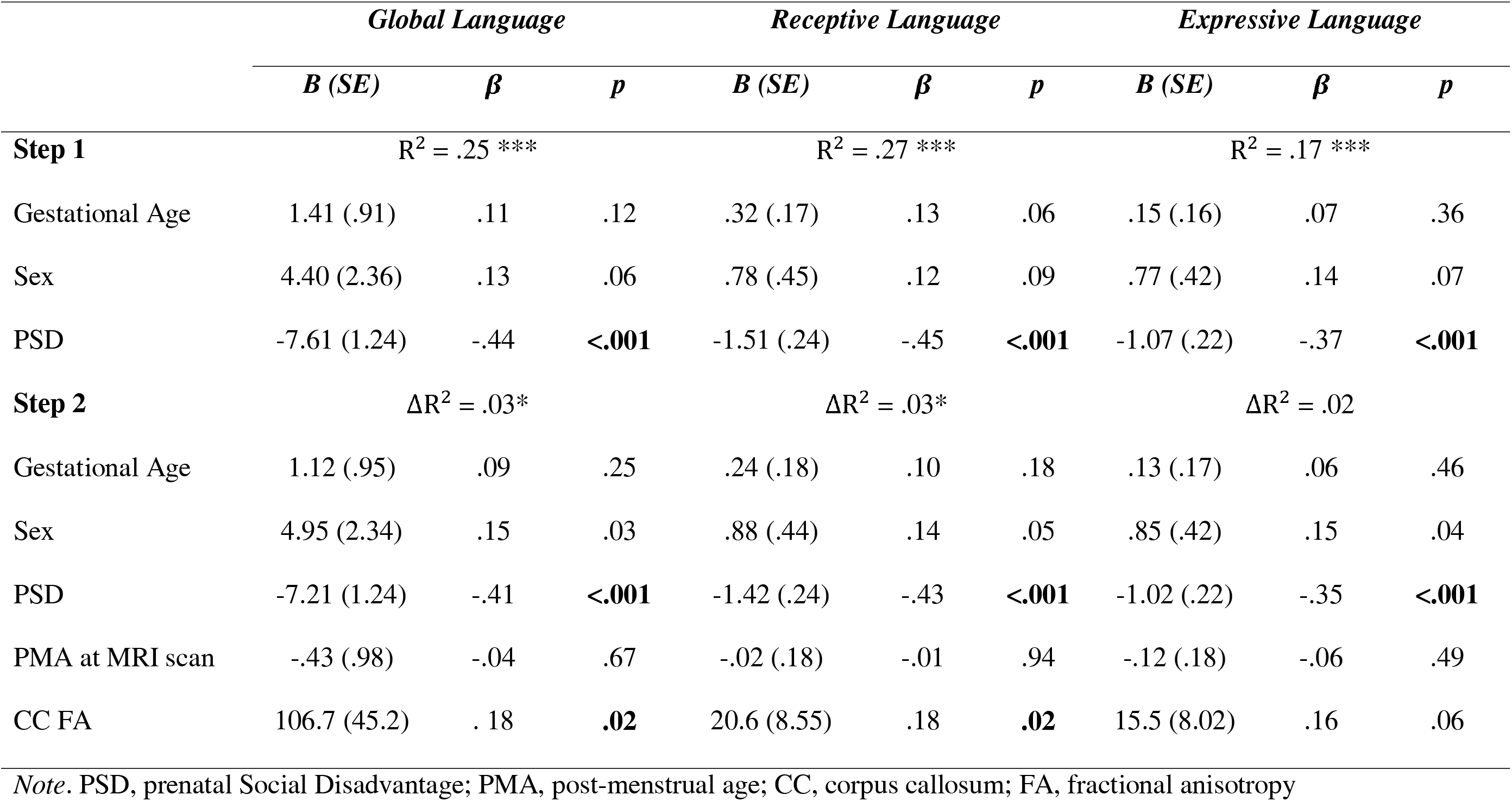
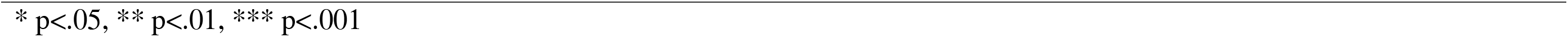
Stepwise associations between prenatal Social Disadvantage, neonatal corpus callosum fractional anisotropy, and language

|  | *Global Language* |  |  | *Receptive Language* |  |  | *Expressive Language* |  |  |
| --- | --- | --- | --- | --- | --- | --- | --- | --- | --- |
| | *B (SE)* | $\beta$ | *p* | *B (SE)* | $\beta$ | *p* | *B (SE)* | $\beta$ | *p* |
| **Step 1** | $R^2 = .25^{***}$ | | | $R^2 = .27^{***}$ | | | $R^2 = .17^{***}$ | | |
| Gestational Age | 1.41 (.91) | .11 | .12 | .32 (.17) | .13 | .06 | .15 (.16) | .07 | .36 |
| Sex | 4.40 (2.36) | .13 | .06 | .78 (.45) | .12 | .09 | .77 (.42) | .14 | .07 |
| PSD | -7.61 (1.24) | -.44 | **<.001** | -1.51 (.24) | -.45 | **<.001** | -1.07 (.22) | -.37 | **<.001** |
| **Step 2** | $\Delta R^2 = .03^*$ | | | $\Delta R^2 = .03^*$ | | | $\Delta R^2 = .02$ | | |
| Gestational Age | 1.12 (.95) | .09 | .25 | .24 (.18) | .10 | .18 | .13 (.17) | .06 | .46 |
| Sex | 4.95 (2.34) | .15 | .03 | .88 (.44) | .14 | .05 | .85 (.42) | .15 | .04 |
| PSD | -7.21 (1.24) | -.41 | **<.001** | -1.42 (.24) | -.43 | **<.001** | -1.02 (.22) | -.35 | **<.001** |
| PMA at MRI scan | -.43 (.98) | -.04 | .67 | -.02 (.18) | -.01 | .94 | -.12 (.18) | -.06 | .49 |
| CC FA | 106.7 (45.2) | .18 | **.02** | 20.6 (8.55) | .18 | **.02** | 15.5 (8.02) | .16 | .06 |

*Note.* PSD, prenatal Social Disadvantage; PMA, post-menstrual age; CC, corpus callosum; FA, fractional anisotropy
\* $p < .05$ , \*\* $p < .01$ , \*\*\* $p < .001$

#### Mediation Analysis

Given that PSD and neonatal CC FA were associated with language outcomes, we examined whether the indirect effect of PSD to language via neonatal CC FA (*i.e.,* the product *ab;* (Hayes & Rockwood, 2017)) attenuated the association between PSD and language outcome. As shown in Table S8, the indirect effects for global (95% CIs: -0.96 – 0.15) and receptive (95% CIs: -0.19 – 0.03) were not significant.

#### Moderation Analysis

As shown in Table S9, the interaction term between PSD and CC FA was not significant for either global (p=.14) or receptive language (p=.13). Moderation analysis was repeated with postnatal year 2 Social Disadvantage, which similarly showed there was no interaction between neonatal CC FA and 2-year Social Disadvantage on global (B=-72.3, SE=43.1, p=.10) or receptive (B=-13.3, SE=8.24, p=.11) language outcomes.

### Maternal Cognitive Ability and Home Enrichment

When examined separately, maternal TOPF scores (r=.54, p<.001) and Stim-Q Toddler Total scores (r=.32, p=.01) were both correlated with global language scores. We also note that neonatal CC FA was not correlated with maternal TOPF scores (r=.19, p>.05), but was positively correlated with later StimQ-Toddler total scores (r=.25, p=.03). When TOPF and StimQ-Toddler total scores were included simultaneously, along with PSD and neonatal CC FA, as independent variables fitted to global language scores, only PSD was independently associated with language outcomes (p<.001, Table 5).

**Table 5.** Associations between prenatal Social Disadvantage, neonatal corpus callosum fractional anisotropy, and global language outcomes accounting for maternal cognitive ability

|  | *B (SE)* | *β* | *p* |
| --- | --- | --- | --- |
| **Global Language** | | $R^2 = .49^{**}$ | |
| Gestational Age | .75 (1.12) | .07 | .50 |
| Child Sex | 5.52 (3.00) | .18 | .07 |
| PSD | -13.1 (2.75) | -.83 | **<.001** |
| PMA at MRI scan | .47 (1.20) | .04 | .80 |
| CC FA | 71.0 (61.9) | .12 | .26 |
| Maternal TOPF | -.20 (.16) | -.22 | .21 |
| StimQ-Toddler Total | -.16 (.24) | -.08 | .49 |

*Note.* PSD, prenatal Social Disadvantage; TOPF, Test of Premorbid Function, PMA, post-menstrual age; CC, corpus callosum; FA, fractional anisotropy
\* $p < .05$ , \*\* $p < .01$ , \*\*\* $p < .001$

### Bayley-III Cognition

Regression models with covariate factors were repeated using Bayley-III Cognition scores as the dependent variable. As shown in Table S10, CC FA (p=.09) and AD (p=.10) were not related to cognitive outcome (Table S10).

### Sensitivity Analyses

When excluding children with Bayley-III Cognition <70, associations between neonatal CC FA and AD and poorer language outcomes were unchanged (Table S11). Study conclusions also remained the same after excluding children born preterm (Table S11).

## DISCUSSION

In this cohort of prenatally-recruited infants enriched for adversity, PSD was associated with poorer global, receptive, and expressive language outcomes, with PSD explaining approximately 10% more of the variance in receptive language development than expressive language development at age 2 years. Prenatal exposure to maternal Psychosocial Stress was not significant after accounting for PSD. We extend previous research to show that prospective associations between Social Disadvantage and language development may begin *in utero* (Fernald et al., 2013; Justice et al., 2019; Nwanne et al., 2022). Postnatal Social Disadvantage was also related to language outcomes, suggesting that ongoing exposure likely continues to shape language development into childhood. Key postnatal mechanisms previously associated with social disadvantage include difficulty accessing high-quality child education, reduced learning materials in the home, or limited parental time to engage in parent-child interactions where language modelling and conversational turn-taking take place (Hirsh-Pasek et al., 2015; Nwanne et al., 2022; Perkins et al., 2013). We considered maternal cognitive ability as a heritable factor and home enrichment as a postnatal environmental factor correlated with language outcomes, but neither were significant over and above PSD. While our findings might highlight the substantial associations between severe disadvantage on neurodevelopment, we note that sample size was smaller for analyses of maternal cognitive ability and home enrichment. Nonetheless, our findings suggest that PSD may begin to shape language development prior to exposure to postnatal mechanisms known to support language acquisition (Duncan, Magnuson, & Votruba-Drzal, 2017; Lean, Paul, Smyser, & Rogers, 2018; Perkins et al., 2013).

Independent of PSD, lower neonatal CC FA was associated with language outcomes. Lower FA may reflect a combination of reduced axonal packing, smaller axonal diameter, less myelination, or overall more immature white matter (Feldman, Yeatman, Lee, Barde, & Gaman-Bean, 2010). Study findings are somewhat in line with Sket et al. (2019), who also found that lower neonatal CC FA was related to worse parent-report ratings of infant language ability at age 12 months whereas maternal income and education were not significant. Differences in study findings may reflect the fact that our cohort was oversampled for dyads from low-income background, we included more aspects of social disadvantage, and assessed language using the gold-standard Bayley-III. The CC is one of the largest and fastest proliferating white matter tracts (Dubois et al., 2014). Additional work using probabilistic tractography has also shown that the interhemispheric pathways connecting language regions travel through the CC (Xiang et al., 2012). Taken together, neonatal alterations in the neural architecture that facilitates the interhemispheric transfer of information may underlie the development of early problems in language processing (Friedrich et al., 2020; Ocklenburg, Ball, Wolf, Genç, & Güntürkün, 2015). We note that in our study, the CC was not related to cognition. We assessed CC microstructure with probabilistic tractography, whereas some previous studies have used tract-based spatial statistics in neonates and most commonly found that microstructure of the genu predicts cognitive development (Duerden et al., 2015; Feng et al., 2019; Parikh et al., 2021). The genu projects into the medial prefrontal cortex, a region known to be important for numerous cognitive functions (van den Heuvel, Mandl, Kahn, & Hulshoff Pol, 2009; Ridderinkhof, Ullsperger, Crone, & Nieuwenhuis, 2004; Simon, Rudebeck, & Rich, 2021). The body and splenium of the CC project into motor and visual regions, respectively, which map onto Brodmann’s semantic function and language processing areas (Ardila, Bernal, & Rosselli, 2016; Hertrich, Dietrich, & Ackermann, 2016; van den Heuvel et al., 2009).

Of particular interest was the potential mediating pathways between PSD and language outcomes via neonatal white matter tracts that subserve language. However, CC microstructure did not mediate the relationship between PSD and language outcomes. Moderation analyses further suggested that the association between CC microstructure and language outcome did not vary as a function of PSD. While previous work in the eLABE cohort has demonstrated that associations between measures of maternal stress and inflammation during pregnancy on neonatal brain macro- and micro-structure depend on the level of PSD (Herzberg et al., 2023; Lean et al., 2022; Sanders et al., 2024), our study results suggest that individual differences in neonatal CC white matter microstructure relate to language ability at age 2 years regardless of the level of PSD. Interactions between neonatal CC white matter and 2-year postnatal Social Disadvantage on language ability were also not significant, suggesting that infants with pronounced neonatal microstructural alterations were not differentially more sensitive to postnatal disadvantage (Miller, Chau, Synnes, Miller, & Grunau, 2022). While the CC is one of the earliest developing fibers, myelination and maturation continues throughout early childhood (Lebel, Walker, Leemans, Phillips, & Beaulieu, 2008). Longer-term alterations in CC development from birth and throughout early life may come to mediate or moderate associations between PSD and language outcomes at older ages. Mechanistic pathways may also be more discernable later in language development when disparities in language ability may have widened as a function ongoing exposure to postnatal social disadvantage. (Armstrong et al., 2017; Fernald et al., 2013; Houweling et al., 2022). This is an important topic for future study.

In contrast to our hypothesis, the uncinate, IFOF, or the SLF/AF bundle were not associated with language outcomes at age 2 years. These tracts have been implicated in language functions in older populations, connecting brain regions in dorsal and ventral language pathways, as well as Wernicke’s and Broca’s areas (Fujii et al., 2016; Ostertag et al., 2023; Urger et al., 2015; Von Der Heide, Skipper, Klobusicky, & Olson, 2013). In the neonate, however, the uncinate, IFOF and SLF/AF have slower rates of myelination than the CC (Horgos et al., 2020; Huang et al., 2006). Nonetheless, findings in two preterm-born cohorts have also shown that neonatal microstructure in the uncinate was not yet related to early language outcomes at age 2 years (Bugada et al., 2021; Dubner et al., 2020). Given that the uncinate, IFOF, and SLF demonstrate protracted trajectories of myelination early childhood into adulthood (Lebel et al., 2008; Olson, Heide, Alm, & Vyas, 2015; Zhang et al., 2007), it may be that later development and/or ongoing alterations in the microstructure of the uncinate, IFOF and SLF/AF will be important for subsequent language outcomes.

Current study findings have critical clinical and research implications. Associations between PSD on language outcomes, independent of maternal cognitive ability, further informs the type of preventions focused on PSD. For example, supplementing income and expanding access to social welfare programs during pregnancy may address some of the modifiable aspects of PSD related to language development in offspring (Goldfeld et al., 2024). Further, future studies might explore the language exposure of the fetus prenatally to investigate whether such exposures during pregnancy might enhance language development in offspring. Families experiencing social adversity during the prenatal period and into the early child rearing years may also benefit from access to government-funded services that provide families with education and/or in-home materials that focus on ways that parents can support infant and toddler language skill acquisition in the context of the parent-child relationship (Health Resource & Service Administration, 2024). Study strengths included longitudinal study design beginning in pregnancy, oversampling of mothers from low-income backgrounds, assessment of multidimensional constructs measuring adversity experienced *in utero*, high quality neonatal dMRI data, standardized task-based language assessments, and consideration of maternal cognitive ability and home enrichment.

Study limitations include technical considerations regarding probabilistic tractography of the SLF and AF (see Supporting Information), which are slow to myelinate and difficult to track individually in the neonate compared to the CC which is larger and more established at birth (Janelle et al., 2022; Liang et al., 2022; Schilling et al., 2022). Additionally, prenatal and postnatal Social Disadvantage were inter-correlated and thus stable over time, potentially reflecting the lack of social mobility in our Midwestern cohort (Connor & Storper, 2020). As such, we were unable to disentangle the possible importance of timing of exposure to Social Disadvantage on language outcomes.

### Conclusions

These findings suggest that PSD is associated with language delays by age 2 years. Lower neonatal CC FA was also related to language delays by age 2 years, independent of PSD. However, a clear mechanistic relationship between PSD and neonatal CC microstructure on language outcome was not found at this early developmental point. Such associations may emerge as language disparities continue to widen over time with ongoing exposure to Social Disadvantage. Nonetheless, findings highlight the CC as an important commissural fiber that may underlie early language development and thus identify infants who may benefit from early referral to language therapies before observable language delays emerge. Importantly, findings also underscore the importance of broadening access to social welfare programs that reduce the burden and stressors of social disadvantage during the critical prenatal window to support infant neurodevelopmental outcomes.

## Data Availability

Data can be made available to qualified researchers by written request to Primary Investigators Drs. Luby, Smyser, and Warner under the guidance of a formal data sharing agreement. Data will also be available via the open access NIMH Data Archive repository at the conclusion of this study.

## Key Points and Relevance

- Prenatal Social Disadvantage (PSD) is associated with early child language delays, independent of maternal cognitive ability and home enrichment.
- After accounting for PSD, lower neonatal corpus callosum fractional anisotropy was associated with language delays by age 2 years.
- Neonatal white matter microstructure did not mediate the link between PSD and language outcomes at age 2 years, suggesting independent pathways to language outcomes.
- Findings reveal white matter correlates to help identify infants who may benefit from early intervention before delays emerge and underscore the importance of broadening access to social welfare programs during the critical prenatal window of neurodevelopment.

## Acknowledgements

This study was supported by the National Institute of Mental Health (R01 MH113883, K01 MH122735), March of Dimes Prematurity Research Center at Washington University, Washington University Intellectual and Developmental Disability Research Center (P50 HD103525), Children’s Discovery Institute, McDonnell Center for Systems Neuroscience, and a NARSAD Young Investigator Grant (No. 28521) from the Brain and Behavior Research Foundation. We thank the families involved in this study.

## Conflicts of Interest

The authors have no real or perceived conflicts of interest to disclose.

## Abbreviations

AD: Axial diffusivity
AF: Arcuate fasciculus
Bayley-III: Bayley Scales of Infant and Toddler Development-III
CC: Corpus callosum
dMRI: Diffusion magnetic resonance imaging
eLABE: Early Life Adversity and Biological Embedding study
FA: Fractional anisotropy
FSL: FMRIB Software Library
GA: Gestational age
IFOF: Inferior fronto-occipital fasciculus
MD: Mean diffusivity
PMA: Postmenstrual age
PSD: Prenatal Social Disadvantage
RD: Radial diffusivity
ROI: Regions-of-Interest
SD: Standard deviation
SLF: Superior longitudinal fasciculus

## Notes

### Competing Interest Statement

The authors have declared no competing interest.

### Author Declarations

The Washington University Institutional Review Board gave ethical approval for this work

