## Supporting Information for "Associations between prenatal adversity and neonatal white matter microstructure on language outcomes at age 2 years"

**Contents:**

**Supplemental Figures**

**Figure S1.** Participant flow from recruitment in the larger eLABE cohort (n=398) to inclusion in current study analysis (n=160)

**Figure S2-4.** Probabilistic tractography of the neonatal superior longitudinal fasciculus (SLF) and arcuate fasciculus (AF)

**Supplemental Tables**

**Table S1.** Pearson correlation coefficients between measures of Social Disadvantage and language outcomes at age 2 years

**Table S2.** Comparison of language ability between children assessed before or after the onset of COVID-19 Pandemic Lockdown in St. Louis (March 16th, 2020)

**Table S3.** Descriptive statistics for neonatal dMRI parameters in tracts of interest

**Table S4.** Associations between neonatal dMRI parameters and receptive language outcomes by age 2 years

**Table S5.** Associations between neonatal dMRI parameters and expressive language outcomes by age 2 years

**Table S6.** Associations between neonatal mean and radial diffusivity and global language outcomes at age 2 years

**Table S7.** Associations between corpus callosum axial diffusivity and language outcomes after accounting for prenatal Social Disadvantage

**Table S8.** Mediation analysis of prenatal Social Disadvantage, neonatal corpus callosum fractional anisotropy and axial diffusivity, and global/receptive language outcomes at age 2 years

**Table S9.** Formal moderation analyses with prenatal Social Disadvantage and neonatal corpus callosum fractional anisotropy

**Table S10.** Associations between corpus callosum fractional anisotropy and axial diffusivity and cognitive outcome at age 2 years

**Table S11.** Summary of regression analyses after excluding children with severe cognitive delay (Bayley-III cognition <70) or children born preterm (<37 weeks gestation)

**Abbreviations:**

| | |
|---|---|
| AD | Axial diffusivity |
| AF | Arcuate fasciculus |
| Bayley-III | Bayley Scales of Infant and Toddler Development-III |
| CC | Corpus callosum |
| dMRI | Diffusion magnetic resonance imaging |
| eLABE | Early Life Adversity and Biological Embedding |
| FA | Fractional anisotropy |
| FSL | FMRIB Software Library |
| GA | Gestational age |
| IFOF | Inferior fronto-occipital fasciculus |
| MD | Mean diffusivity |
| PMA | Postmenstrual age |
| PSD | Prenatal Social Disadvantage |
| RD | Radial diffusivity |
| ROI | Regions-of-Interest |
| SD | Standard deviation |
| SLF | Superior longitudinal fasciculus |

### SUPPLEMENTAL FIGURES

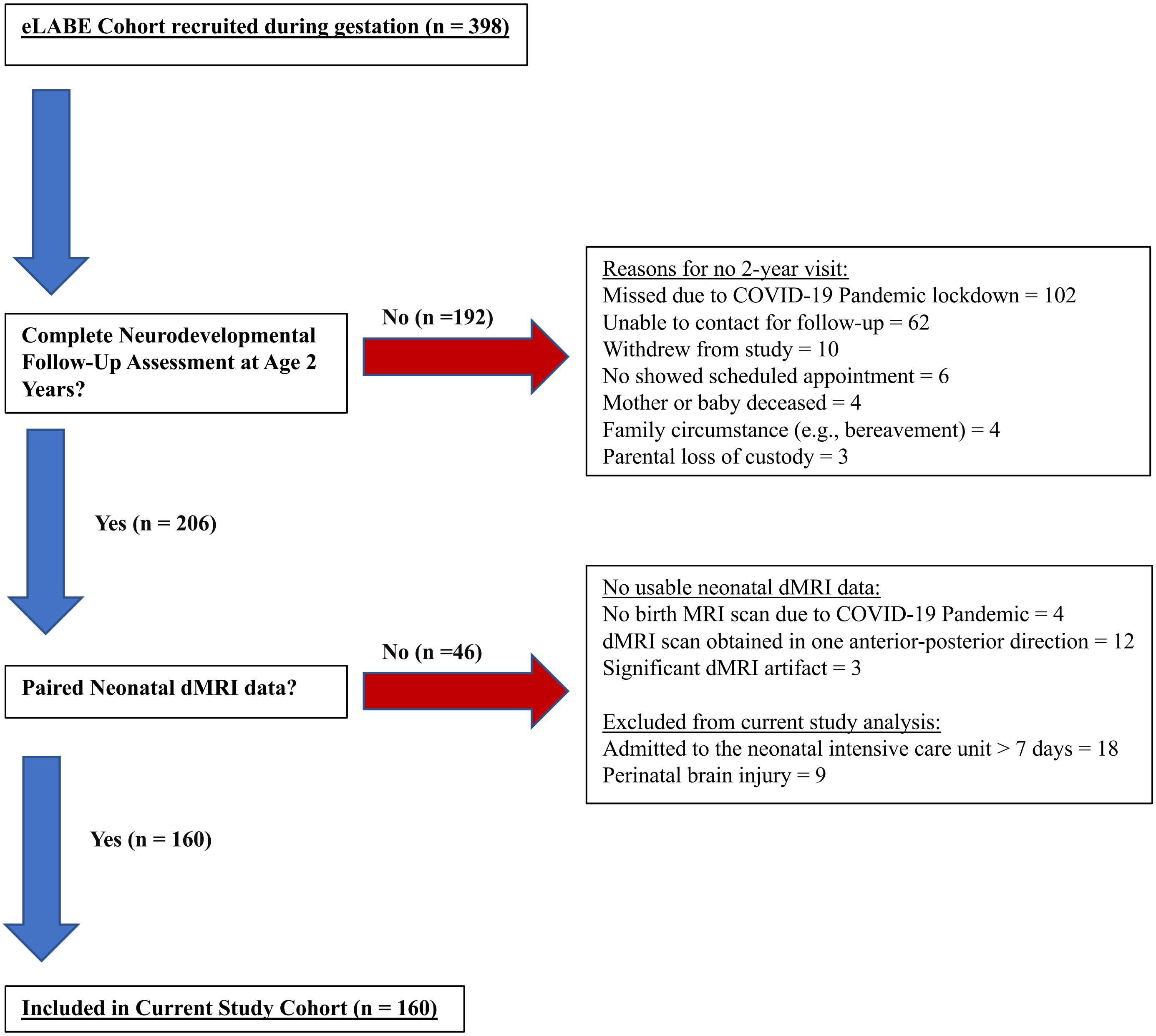


**Figure S1.** Participant flow from recruitment in the larger eLABE cohort (n=398) to inclusion in current study analysis (n=160)

**Neonatal Superior Longitudinal Fasciculus (SLF) and Arcuate Fasciculus (AF)**

The SLF is a superior anterior-posterior tract comprised of three subdivisions that connect temporal, parietal, and frontal brain regions and form part of the dorsal language stream (Fujii et al., 2016; Janelle, Iorio-Morin, D'amour, & Fortin, 2022; Urger et al., 2015). An additional deeper fiber bundle that connects the caudal part of the superior temporal gyrus with the lateral prefrontal cortex in the dorsal language stream has been identified as the AF. The SLF and AF have been linked with similar language abilities including expressive language skills and phonological processing (Fujii et al., 2016; Urger et al., 2015).

In neonates, white matter matures in an anterior-posterior manner, with the SLF being a later developing tract characterized by slower rates of myelination compared to other more established fibers such as the corpus callosum (Zhang et al., 2006). Given the relative immaturity of the SLF in neonates as well as its proximity to the AF, previous studies have shown that reliable reconstruction of the early SLF using tractography is difficult (Janelle et al., 2022; Liang et al., 2022; Schilling et al., 2022). In the current study, we initially undertook probabilistic tractography of the SLF and AF separately but found that fibers from the AF transversed through the same voxel as the SLF (Figure S2).

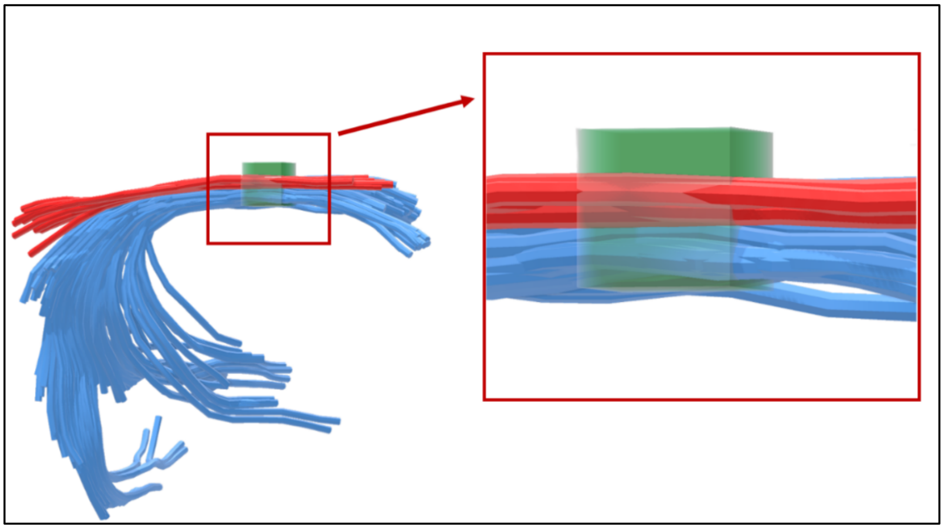

**Figure S2.** Illustration of the superior longitudinal fasciculus (SLF, red) and the arcuate fasciculus (AF, blue) from a representative study neonate. As shown, the SLF and AF transverse through the same 1.75mm$^3$ voxel (green). Tracts and voxel are shown to scale.

Figure S3 shows the seed regions-of-interest (ROIs) on FA maps in a representative study neonate that were used to generate the SLF (A) and the AF (B) using probabilistic tractography. Importantly, Panel C shows the apparent overlap between the outputted SLF and AF (C). We note that adjustment of curvature thresholds (see also Figure S4) or use of exclusion masks did not improve the parcellation of the SLF and AF. Due to these technical issues in delineating the SLF and AF as separate fiber pathways, we acknowledge our probabilistic tractography of the SLF likely contains fibers from the AF. Therefore, for the purposes of the current study, we refer to this white matter pathway as a combined SLF/AF language bundle.

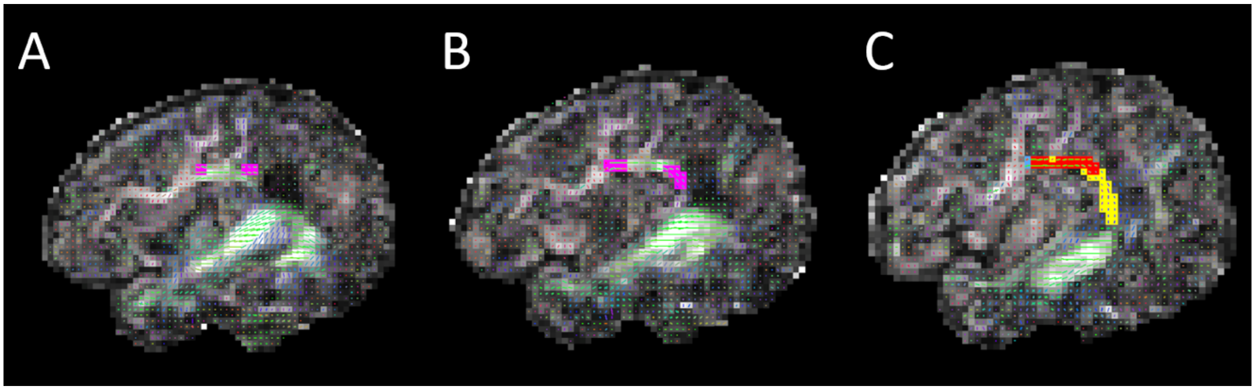


**Figure S3.** Slices of the neonatal brain showing seeds placed at well-defined anatomical features for the superior longitudinal fasciculus (SLF, A) and the arcuate fasciculus (AF, B), and the outputted tracts after performing probabilistic tractography in FSL (C; SLF, blue; AF, yellow; overlapping portions of the SLF and AF, orange).

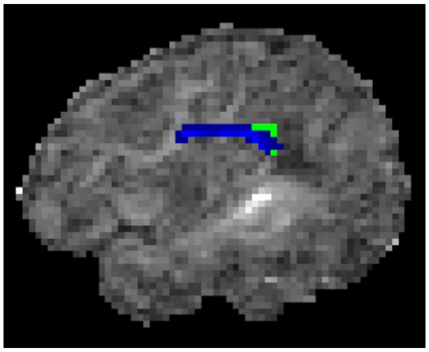

**Figure S4.** Outputted AF after thresholding to retain the streamlines with probability values indicating greater certainty of white matter. As shown, the superior portion of the AF was more readily detected in a representative neonate than the inferior portion of the AF which was not retained after thresholding. The AF is incompletely myelinated and does not fully connect language regions in newborns (Dubois et al., 2014). Therefore, our combined SLF/AF bundle (Figures S2-3) did not include the inferior portion of the AF.

### SUPPLEMENTAL TABLES

#### Pre- and Postnatal Social Disadvantage

Similar to the prenatal timepoint, a latent construct for Social Disadvantage was created using structural equation modeling at age 2 years. This latent factor was based upon the highest level of maternal education, income-to-needs ratio, and area deprivation index percentiles (computed as the average percentile across 16, 20, and 24 months). Social Disadvantage at the prenatal and year 2 timepoints were very highly correlated ($r = .93$, $p < .001$). As expected, Social Disadvantage at age 2 years was inversely correlated with global ($r = -.44$, $p<.001$), receptive ($r=-.44$, $p<.001$), and expressive ($r=-.44$, $p<.001$) language scores at age 2 years. Given the high correlation between prenatal and year 2 social disadvantage likely reflecting a lack of social mobility in our Midwestern sample (Connor & Storper, 2020), it is difficult to distinguish prenatal vs. postnatal influences on language development by age 2 years. It is possible that the influence of social adversity begins prenatally and continues postnatally in our sample.

Please also see Main Text for reporting of formal moderation analysis performed with Social Disadvantage at age 2 years to account for postanal exposure on language outcomes, with similar null results as moderation analysis performed with PSD.

**Table S1.** Pearson correlation coefficients between measures of Social Disadvantage and language outcomes

| | **PSD** | **Year 2 Social Disadvantage** | **Global Language** | **Receptive Language** |
|---|---|---|---|---|
| PSD | - | | | |
| Year 2 Social Disadvantage | .93*** | - | | |
| Global Language | -.47*** | -.44*** | - | |
| Receptive Language | -.49*** | -.44*** | .95*** | - |
| Expressive Language | -.39*** | -.38*** | .93*** | .77*** |

*Note.* PSD, prenatal Social Disadvantage

*p<.05, **p<.01, ***p<.001

**No Differences in Language Scores between Participants who Attended Follow-Up at Age 2 Years Before/After the Onset of COVID-19 Pandemic.**

Recent research has shown that the COVID-19 Pandemic may be associated with difficulties in language development. For example, COVID-19 Pandemic lockdown stay-at-home orders and ongoing safety procedures such as social distancing and school closures may be thought to reduce the number of words that children heard and reduce access to early intervention services (Charney, Camarata, & Chern, 2021; Whitley, Beauchamp, & Brown, 2021). To explore potential differences in language ability between children who completed in-person developmental assessments before (n = 29) and after (n = 131) the onset of COVID-19 Pandemic lockdown in St. Louis (March 16$^{th}$, 2020), differences in mean global, receptive, and expressive language scores were compared using independent samples t-tests and effect sizes calculated using Cohen's *d*. As shown in Table S2 below, there were no statistical differences in language development and effect sizes were relatively small-to-modest between children assessed pre- and post the COVID-19 Pandemic Lockdown.

**Table S2.** Comparison of language ability between children assessed before or after the onset of COVID-19 Pandemic Lockdown in St. Louis (March 16$^{th}$, 2020)

| **M (SD)** | **Pre-Lockdown (n = 29)** | **Post-Lockdown (n = 131)** | ***t*** | ***p*** | ***d*** |
|---|---|---|---|---|---|
| Global language | 91.6 (15.2) | 89.1 (17.3) | .72 | .50 | .15 |
| Receptive language | 8.2 (3.1) | 7.5 (3.3) | 1.14 | .24 | .24 |
| Expressive language | 8.8 (2.4) | 8.8 (2.9) | .11 | .98 | .03 |

**Table S3.** Descriptive statistics for neonatal dMRI parameters in tracts of interest

| | ***FA*** | | | ***MD*** | | | ***AD*** | | | ***RD*** | | |
|---|---|---|---|---|---|---|---|---|---|---|---|---|
| | ***X*** | ***SD*** | **Range** | ***X*** | ***SD*** | **Range** | ***X*** | ***SD*** | **Range** | ***X*** | ***SD*** | **Range** |
| **CC** | .341 | .029 | (.252 - .411) | 1.006 | .033 | (.929 – 1.108) | 1.399 | .028 | (1.278 – 1.466) | .809 | .044 | (.712 - .931) |
| **R SLF/AF** | .158 | .030 | (.077 - .241) | 1.646 | .157 | (1.311–2.134) | 1.873 | .136 | (1.597 – 2.286) | 1.533 | .169 | (1.150 – 2.057) |
| **L SLF/AF** | .169 | .031 | (.068 - .246) | 1.599 | .144 | (1.314 – 2.165) | 1.832 | .121 | (1.591 – 2.294) | 1.482 | .157 | (1.151 – 2.100) |
| **R Uncinate** | .230 | .026 | (.143 - .300) | 1.063 | .043 | (.937 – 1.186) | 1.331 | .044 | (1.177 – 1.469) | .929 | .050 | (.801 – 1.096) |
| **L Uncinate** | .233 | .026 | (.143 - .302) | 1.043 | .043 | (.916 – 1.136) | 1.311 | .053 | (1.131 – 1.458) | .908 | .048 | (.692 – 1.040) |
| **R IFOF** | .232 | .035 | (.144 – .315) | 1.094 | .054 | (.965 – 1.259) | 1.361 | .034 | (1.252 – 1.445) | .961 | .068 | (.821 – 1.166) |
| **L IFOF** | .231 | .034 | (.149 - .306) | 1.092 | .051 | (.966 – 1.197) | 1.358 | .035 | (1.233 – 1.424) | .959 | .064 | (.831 – 1.099) |

*Note.* FA, fractional anisotropy; MD, mean diffusivity; AD, axial diffusivity; RD, radial diffusivity; CC, corpus callosum; SLF/AF, superior longitudinal fasciculus/arcuate fasciculus; IFOF, inferior fronto-occipital fasciculus, R, right; L, left

**Table S4.** Associations between neonatal dMRI parameters and receptive language outcomes by age 2 years

| | *FA* | | | | *MD* | | | | *AD* | | | | *RD* | | | |
|---|---|---|---|---|---|---|---|---|---|---|---|---|---|---|---|---|
| | ***B (SE)*** | ***β*** | ***p*** | ***q*** | ***B (SE)*** | ***β*** | ***p*** | ***q*** | ***B (SE)*** | ***β*** | ***p*** | ***q*** | ***B (SE)*** | ***β*** | ***p*** | ***q*** |
| **Model 1: Corpus Callosum** | | | | | | | | | | | | | | | | |
| | $R^2$ = .13*** | | | | $R^2$ = .09** | | | | $R^2$ = .12*** | | | | $R^2$ = .11** | | | |
| GA | .43 (.20) | .18 | .03 | .04 | .48 (.20) | .20 | .02 | .03 | .39 (.20) | .16 | .05 | .05 | .47 (.20) | .19 | .02 | .04 |
| Sex | 1.08 (.49) | .17 | .03 | .06 | .94 (.50) | .15 | .06 | .07 | 1.12 (.50) | .17 | .03 | .08 | .98 (.50) | .15 | .05 | .07 |
| PMA at scan | .03 (.21) | .01 | .90 | .89 | .15 (.23) | .06 | .52 | .52 | .39 (.20) | .16 | .05 | .24 | .05 (.22) | .02 | .82 | .82 |
| **CC FA** | **26.5 (9.41)** | **.24** | **.01** | **.01** | -8.54 (9.01) | -.09 | .35 | .34 | **21.0 (9.08)** | **.18** | **.02** | **.02** | -12.5 (6.62) | -.17 | .06 | .06 |
| **Model 2: R SLF/AF** | | | | | | | | | | | | | | | | |
| | $R^2$ = .09** | | | | $R^2$ = .09** | | | | $R^2$ = .09** | | | | $R^2$ = .09** | | | |
| GA | .46 (.20) | .19 | .03 | .04 | .44 (.20) | .18 | .03 | .03 | .44 (.20) | .18 | .03 | .04 | .44 (.20) | .18 | .03 | .04 |
| Sex | .95 (.50) | .15 | .06 | .06 | .91 (.50) | .14 | .07 | .07 | .90 (.50) | .14 | .07 | .08 | .91 (.50) | .14 | .07 | .07 |
| PMA at scan | .22 (.21) | .09 | .30 | .55 | .16 (.21) | .07 | .44 | .50 | .16 (.21) | .07 | .44 | .44 | .17 (.21) | .07 | .43 | .55 |
| R SLF/AF | 4.76 (9.33) | .05 | .61 | .61 | -2.00 (1.81) | -.10 | .27 | .27 | -2.49 (2.08) | -.10 | .23 | .23 | -1.80 (.169) | -.09 | .29 | .29 |
| **Model 3: L SLF /AF** | | | | | | | | | | | | | | | | |
| | $R^2$ = .09** | | | | $R^2$ = .09** | | | | $R^2$ = .09** | | | | $R^2$ = .09** | | | |
| GA | .46 (.20) | .19 | .03 | .04 | .46 (.20) | .19 | .03 | .03 | .46 (.20) | .19 | .02 | .04 | .46 (.20) | .19 | .03 | .04 |
| Sex | .95 (.50) | .15 | .06 | .06 | .95 (.50) | .15 | .06 | .07 | .95 (.50) | .15 | .06 | .08 | .95 (.50) | .15 | .06 | .07 |
| PMA at scan | .23 (.22) | .10 | .31 | .55 | .22 (.22) | .09 | .32 | .50 | .23 (.21) | .10 | .29 | .39 | .21 (.22) | .09 | .34 | .55 |
| L SLF/AF | 3.04 (9.73) | .03 | .76 | .76 | -.89 (2.03) | -.04 | .66 | .66 | -.82 (2.38) | -.03 | .73 | .73 | -.88 (1.88) | -.04 | .64 | .64 |

**Table S4.** Associations between neonatal dMRI parameters and receptive language outcomes by age 2 years

| | ***FA*** | | | | ***MD*** | | | | ***AD*** | | | | ***RD*** | | | |
|---|---|---|---|---|---|---|---|---|---|---|---|---|---|---|---|---|
| | ***B (SE)*** | ***β*** | ***p*** | ***q*** | ***B (SE)*** | ***β*** | ***p*** | ***q*** | ***B (SE)*** | ***β*** | ***p*** | ***q*** | ***B (SE)*** | ***β*** | ***p*** | ***q*** |
| **Model 4: R Uncinate** | | | | | | | | | | | | | | | | |
| | $R^2$ = .08** | | | | $R^2$ = .08** | | | | $R^2$ = .09** | | | | $R^2$ = .09** | | | |
| GA | .47 (.20) | .20 | .02 | .04 | .49 (.20) | .20 | .02 | .03 | .50 (.20) | .21 | .01 | .04 | .48 (.20) | .20 | .02 | .04 |
| Sex | .95 (.50) | .15 | .06 | .06 | .92 (.50) | .14 | .07 | .07 | .90 (.50) | .14 | .08 | .08 | .94 (.50) | .14 | .06 | .07 |
| PMA at scan | .28 (.21) | .12 | .19 | .55 | .17 (.22) | .07 | .43 | .50 | .21 (.20) | .09 | .30 | .39 | .19 (.22) | .08 | .40 | .55 |
| R Uncinate | -2.35 (10.7) | -.02 | .83 | .83 | -5.53 (6.51) | -.07 | .40 | .40 | -5.62 (5.90) | -.08 | .34 | .34 | -3.84 (5.79) | -.06 | .51 | .51 |
| **Model 5: L Uncinate** | | | | | | | | | | | | | | | | |
| | $R^2$ = .09** | | | | $R^2$ = .09** | | | | $R^2$ = .08** | | | | $R^2$ = .08** | | | |
| GA | .49 (.20) | .20 | .02 | .04 | .47 (.20) | .19 | .02 | .03 | .47 (.20) | .19 | .02 | .04 | .47 (.20) | .19 | .02 | .04 |
| Sex | .94 (.50) | .15 | .06 | .06 | .95 (.50) | .15 | .06 | .07 | .95 (.50) | .15 | .06 | .08 | .95 (.50) | .15 | .06 | .07 |
| PMA at scan | .29 (.21) | .12 | .16 | .55 | .28 (.21) | .12 | .20 | .50 | .26 (.20) | .11 | .20 | .39 | .26 (.22) | .11 | .23 | .55 |
| L Uncinate | -4.73 (10.7) | -.04 | .66 | .66 | 1.34 (6.48) | .02 | .84 | .84 | -.07 (4.91) | -.00 | .99 | .99 | .25 (5.88) | .00 | .97 | .97 |
| **Model 6: R IFOF** | | | | | | | | | | | | | | | | |
| | $R^2$ = .09** | | | | $R^2$ = .09** | | | | $R^2$ = .10** | | | | $R^2$ = .10** | | | |
| GA | .46 (.20) | .19 | .03 | .04 | .48 (.20) | .20 | .02 | .03 | .44 (.20) | .18 | .03 | .04 | .48 (.20) | .20 | .02 | .04 |
| Sex | .96 (.50) | .15 | .06 | .06 | .98 (.50) | .15 | .05 | .07 | 1.08 (.50) | .17 | .03 | .08 | .96 (.50) | .15 | .06 | .07 |

**Table S4.** Associations between neonatal dMRI parameters and receptive language outcomes by age 2 years

| | ***FA*** | | | | ***MD*** | | | | ***AD*** | | | | ***RD*** | | | |
|---|---|---|---|---|---|---|---|---|---|---|---|---|---|---|---|---|
| | ***B (SE)*** | ***β*** | ***p*** | ***q*** | ***B (SE)*** | ***β*** | ***p*** | ***q*** | ***B (SE)*** | ***β*** | ***p*** | ***q*** | ***B (SE)*** | ***β*** | ***p*** | ***q*** |
| PMA at scan | .16 (.24) | .07 | .50 | .68 | .37 (.24) | .16 | .13 | .40 | .41 (.21) | .18 | .05 | .24 | .31 (.25) | .13 | .21 | .55 |
| R IFOF | 6.84 (9.16) | .07 | .46 | .46 | 4.37 (6.02) | .07 | .47 | .47 | 14.9 (8.01) | .16 | .07 | .07 | 1.58 (4.83) | .03 | .74 | .74 |
| **Model 7: L IFOF** | | | | | | | | | | | | | | | | |
| | $R^2 = .08^{**}$ | | | | $R^2 = .09^{**}$ | | | | $R^2 = .09^{**}$ | | | | $R^2 = .09^{**}$ | | | |
| GA | .47 (.20) | .19 | .02 | .04 | .47 (.20) | .19 | .02 | .03 | .44 (.20) | .18 | .03 | .04 | .47 (.20) | .20 | .02 | .04 |
| Sex | .95 (.50) | .15 | .06 | .06 | .97 (.50) | .15 | .05 | .07 | 1.05 (.51) | .16 | .04 | .08 | .95 (.50) | .15 | .06 | .07 |
| PMA at scan | .27 (.23) | .11 | .26 | .55 | .40 (.25) | .17 | .11 | .50 | .36 (.21) | .15 | .09 | .28 | .37 (.25) | .16 | .14 | .55 |
| L IFOF | -.45 (9.35) | -.01 | .96 | .96 | 5.77 (6.44) | .09 | .37 | .37 | 9.04 (8.05) | .10 | .26 | .27 | 3.65 (5.13) | .07 | .48 | .48 |
| **Model 8: R CST** | | | | | | | | | | | | | | | | |
| | $R^2 = .09^{**}$ | | | | $R^2 = .09^{**}$ | | | | $R^2 = .09^{**}$ | | | | $R^2 = .09^{**}$ | | | |
| GA | .42 (.20) | .18 | .04 | .04 | .44 (.20) | .18 | .03 | .03 | .45 (.20) | .19 | .03 | .04 | .43 (.20) | .18 | .04 | .04 |
| Sex | .97 (.50) | .15 | .06 | .06 | .94 (.50) | .15 | .06 | .07 | .94 (.50) | .15 | .06 | .08 | .95 (.50) | .15 | .06 | .07 |
| PMA at scan | .14 (.22) | .06 | .53 | .68 | .17 (.22) | .07 | .45 | .50 | .20 (.22) | .09 | .35 | .39 | .15 (.22) | .06 | .49 | .55 |
| R CST | 20.8 (16.0) | .12 | .19 | .19 | -3.03 (3.14) | -.09 | .34 | .34 | -2.16 (3.57) | -.05 | .55 | .55 | -3.24 (2.92) | -.10 | .27 | .27 |
| **Model 9: L CST** | | | | | | | | | | | | | | | | |
| | $R^2 = .10^{**}$ | | | | $R^2 = .09^{**}$ | | | | $R^2 = .08^{**}$ | | | | $R^2 = .08^{**}$ | | | |
| GA | .42 (.20) | .17 | .04 | .04 | .45 (.20) | .18 | .03 | .03 | .46 (.20) | .19 | .03 | .04 | .44 (.20) | .18 | .03 | .04 |
| Sex | 1.01 (.50) | .16 | .05 | .06 | .95 (.50) | .15 | .06 | .07 | .95 (.50) | .15 | .06 | .08 | .95 (.50) | .15 | .06 | .07 |

**Table S4.** Associations between neonatal dMRI parameters and receptive language outcomes by age 2 years

| | ***FA*** | | | | ***MD*** | | | | ***AD*** | | | | ***RD*** | | | |
|---|---|---|---|---|---|---|---|---|---|---|---|---|---|---|---|---|
| | ***B (SE)*** | ***β*** | ***p*** | ***q*** | ***B (SE)*** | ***β*** | ***p*** | ***q*** | ***B (SE)*** | ***β*** | ***p*** | ***q*** | ***B (SE)*** | ***β*** | ***p*** | ***q*** |
| PMA at scan | .11 (.22) | .05 | .61 | .69 | .19 (.22) | .08 | .40 | .50 | .24 (.22) | .10 | .28 | .39 | .17 (.22) | .07 | .46 | .55 |
| L CST | 22.2 (14.9) | .14 | .14 | .14 | -2.27 (3.19) | -.07 | .48 | .48 | -.85 (3.65) | -.02 | -.23 | .82 | -2.66 (2.96) | -.08 | .37 | .37 |

*Note.* FA, fractional anisotropy; MD, mean diffusivity; AD, axial diffusivity; RD, radial diffusivity; GA, gestational age; PMA, postmenstrual age; SLF/AF, superior longitudinal fasciculus/arcuate fasciculus; IFOF, inferior fronto-occipital fasciculus; CST, corticospinal tract; R, right; L, left

*p<.05, **p<.01, ***p<.001

**Table S5.** Associations between neonatal dMRI parameters and expressive language outcomes by age 2 years

| | ***FA*** | | | | ***MD*** | | | | ***AD*** | | | | ***RD*** | | | |
|---|---|---|---|---|---|---|---|---|---|---|---|---|---|---|---|---|
| | ***B (SE)*** | ***β*** | ***p*** | ***q*** | ***B (SE)*** | ***β*** | ***p*** | ***q*** | ***B (SE)*** | ***β*** | ***p*** | ***q*** | ***B (SE)*** | ***β*** | ***p*** | ***q*** |
| **Model 1: Corpus Callosum** | | | | | | | | | | | | | | | | |
| | $R^2 = .08$** | | | | $R^2 = .05$ | | | | $R^2 = .07$* | | | | $R^2 = .05$ | | | |
| GA | .26 (.18) | .12 | .15 | .15 | .30 (.18) | .14 | .10 | .12 | .23 (.18) | .11 | .20 | .20 | .29 (.18) | .14 | .11 | .11 |
| Sex | .99 (.44) | .17 | .03 | .05 | .89 (.45) | .16 | .05 | .05 | 1.01 (.45) | .18 | .03 | .06 | .91 (.44) | .16 | .04 | .05 |
| PMA at scan | -.09 (.19) | -.04 | .63 | .84 | -.00 (.20) | -.00 | .99 | .99 | .18 (.18) | .09 | .33 | .76 | -.07 (.20) | -.04 | .71 | .80 |
| **CC FA** | **19.8 (8.47)** | **.20** | **.02** | **.02** | -6.58 (8.05) | -.08 | .41 | .41 | 15.3 (8.15) | .15 | .06 | .06 | -9.44 (5.93) | -.15 | .11 | .11 |
| **Model 2: R SLF/AF** | | | | | | | | | | | | | | | | |
| | $R^2 = .05$ | | | | $R^2 = .05$ | | | | $R^2 = .05$ | | | | $R^2 = .05$ | | | |
| GA | .31 (.18) | .15 | .09 | .15 | .29 (.18) | .14 | .11 | .12 | .29 (.18) | .13 | .12 | .18 | .29 (.18) | .14 | .11 | .11 |
| Sex | .90 (.45) | .16 | .05 | .05 | .89 (.45) | .16 | .05 | .05 | .89 (.45) | .16 | .05 | .06 | .90 (.45) | .16 | .05 | .05 |
| PMA at scan | .13 (.19) | .07 | .48 | .84 | .08 (.19) | .04 | .66 | .85 | .07 (.19) | .04 | .69 | .76 | .09 (.19) | .04 | .65 | .80 |
| R SLF/AF | -5.74 (8.33) | -.06 | .49 | .49 | .00 (1.63) | .00 | .99 | .99 | -.24 (1.86) | -.01 | .90 | .90 | .09 (1.52) | .01 | .96 | .96 |
| **Model 3: L SLF /AF** | | | | | | | | | | | | | | | | |
| | $R^2 = .05$ | | | | $R^2 = .05$ | | | | $R^2 = .05$ | | | | $R^2 = .05$ | | | |
| GA | .29 (.19) | .14 | .11 | .15 | .29 (.18) | .14 | .11 | .12 | .29 (.18) | .14 | .11 | .18 | .29 (.18) | .14 | .11 | .11 |
| Sex | .89 (.45) | .16 | .05 | .05 | .89 (.45) | .16 | .05 | .05 | .90 (.45) | .16 | .05 | .06 | .89 (.45) | .16 | .05 | .05 |

**Table S5.** Associations between neonatal dMRI parameters and expressive language outcomes by age 2 years

| | ***FA*** | | | | ***MD*** | | | | ***AD*** | | | | ***RD*** | | | |
|---|---|---|---|---|---|---|---|---|---|---|---|---|---|---|---|---|
| | ***B (SE)*** | ***β*** | ***p*** | ***q*** | ***B (SE)*** | ***β*** | ***p*** | ***q*** | ***B (SE)*** | ***β*** | ***p*** | ***q*** | ***B (SE)*** | ***β*** | ***p*** | ***q*** |
| PMA at scan | .09 (.20) | .04 | .65 | .84 | .09 (.20) | .04 | .65 | .85 | .10 (.19) | .05 | .60 | .76 | .08 (.20) | .04 | .67 | .80 |
| L SLF/AF | -.53 (8.69) | -.01 | .95 | .95 | .10 (1.82) | .01 | .96 | .95 | .46 (2.13) | .02 | .83 | .83 | -.01 (1.68) | -.01 | .99 | .99 |
| **Model 4: R Uncinate** | | | | | | | | | | | | | | | | |
| | $R^2 = .05$ | | | | $R^2 = .05$ | | | | $R^2 = .05$ | | | | $R^2 = .05$ | | | |
| GA | .29 (.18) | .14 | .11 | .15 | .30 (.18) | .14 | .10 | .12 | .31 (.18) | .14 | .09 | .18 | .30 (.18) | .14 | .10 | .11 |
| Sex | .89 (.45) | .16 | .05 | .05 | .87 (.45) | .15 | .05 | .05 | .86 (.45) | .15 | .06 | .06 | .88 (.45) | .16 | .05 | .05 |
| PMA at scan | .09 (.19) | .05 | .63 | .84 | .03 (.20) | .02 | .88 | .98 | .06 (.18) | .03 | .76 | .76 | .04 (.20) | .02 | .86 | .85 |
| R Uncinate | -1.13 (9.51) | -.01 | .91 | .91 | -3.38 (5.82) | -.05 | .56 | .56 | -3.11 (5.28) | -.05 | .56 | .56 | -2.51 (5.17) | -.04 | .63 | .63 |
| **Model 5: L Uncinate** | | | | | | | | | | | | | | | | |
| | $R^2 = .05$ | | | | $R^2 = .05$ | | | | $R^2 = .05$ | | | | $R^2 = .08$** | | | |
| GA | .30 (.18) | .14 | .10 | .15 | .28 (.18) | .13 | .12 | .12 | .27 (.18) | .13 | .14 | .18 | .29 (.18) | .14 | .11 | .11 |
| Sex | .89 (.45) | .15 | .05 | .05 | .88 (.45) | .16 | .05 | .05 | .90 (.45) | .16 | .05 | .06 | .89 (.45) | .16 | .05 | .05 |
| PMA at scan | .11 (.18) | .05 | .56 | .84 | .15 (.19) | .08 | .42 | .85 | .11 (.18) | .05 | .53 | .76 | .15 (.19) | .07 | .45 | .80 |
| L Uncinate | -3.58 (9.53) | -.03 | .71 | .71 | 4.89 (5.77) | .07 | .40 | .40 | 2.85 (4.38) | .05 | .52 | .52 | 3.79 (5.24) | .06 | .47 | .47 |
| **Model 6: R IFOF** | | | | | | | | | | | | | | | | |
| | $R^2 = .05$ | | | | $R^2 = .05$ | | | | $R^2 = .05$ | | | | $R^2 = .05$ | | | |

**Table S5.** Associations between neonatal dMRI parameters and expressive language outcomes by age 2 years

| | ***FA*** | | | | ***MD*** | | | | ***AD*** | | | | ***RD*** | | | |
|---|---|---|---|---|---|---|---|---|---|---|---|---|---|---|---|---|
| | ***B (SE)*** | ***β*** | ***p*** | ***q*** | ***B (SE)*** | ***β*** | ***p*** | ***q*** | ***B (SE)*** | ***β*** | ***p*** | ***q*** | ***B (SE)*** | ***β*** | ***p*** | ***q*** |
| GA | .29 (.18) | .14 | .11 | .15 | .30 (.18) | .14 | .10 | .12 | .26 (.18) | .12 | .14 | .18 | .30 (.18) | .14 | .10 | .11 |
| Sex | .89 (.45) | .16 | .05 | .05 | .93 (.45) | .16 | .04 | .05 | 1.00 (.45) | .18 | .03 | .06 | .92 (.45) | .16 | .04 | .05 |
| PMA at scan | .10 (.21) | .05 | .64 | .84 | .24 (.22) | .12 | .27 | .85 | .21 (.19) | .10 | .27 | .76 | .21 (.22) | .10 | .34 | .80 |
| R IFOF | -1.97 (8.19) | -.01 | .90 | .90 | 6.38 (5.35) | .12 | .24 | .24 | 12.2 (7.17) | .15 | .09 | .09 | 3.99 (4.30) | .10 | .36 | .36 |
| **Model 7: L IFOF** | | | | | | | | | | | | | | | | |
| | $R^2 = .05$ | | | | $R^2 = .05$ | | | | $R^2 = .06$* | | | | $R^2 = .09$** | | | |
| GA | .29 (.18) | .14 | .11 | .15 | .29 (.18) | .13 | .11 | .12 | .25 (.18) | .12 | .16 | .18 | .29 (.18) | .14 | .11 | .11 |
| Sex | .89 (.45) | .16 | .05 | .05 | .92 (.45) | .16 | .04 | .05 | 1.02 (.45) | .18 | .03 | .06 | .89 (.45) | .16 | .05 | .05 |
| PMA at scan | .10 (.21) | .05 | .63 | .84 | .25 (.22) | .12 | .26 | .85 | .22 (.19) | .11 | .26 | .76 | .21 (.22) | .10 | .34 | .80 |
| L IFOF | -1.21 (8.35) | -.01 | .89 | .89 | 6.91 (5.74) | .12 | .23 | .23 | 11.6 (7.16) | .14 | .11 | .11 | 4.22 (4.58) | .10 | .36 | .36 |
| **Model 8: R CST** | | | | | | | | | | | | | | | | |
| | $R^2 = .05$ | | | | $R^2 = .05$ | | | | $R^2 = .05$ | | | | $R^2 = .05$ | | | |
| GA | .28 (.18) | .13 | .13 | .15 | .30 (.18) | .14 | .10 | .12 | .31 (.18) | .15 | .09 | .18 | .30 (.18) | .14 | .10 | .11 |
| Sex | .90 (.45) | .16 | .05 | .05 | .90 (.45) | .16 | .05 | .05 | .91 (.45) | .16 | .05 | .06 | .90 (.45) | .16 | .05 | .05 |
| PMA at scan | .05 (.19) | .02 | .80 | .90 | .12 (.19) | .06 | .53 | .85 | .15 (.19) | .08 | .43 | .76 | .11 (.19) | .05 | .58 | .80 |
| R CST | 5.92 (14.3) | .04 | .68 | .68 | 1.23 (2.81) | .04 | .66 | .66 | 2.67 (3.19) | .08 | .40 | .40 | .71 (2.62) | .03 | .79 | .79 |

**Table S5.** Associations between neonatal dMRI parameters and expressive language outcomes by age 2 years

| | ***FA*** | | | | ***MD*** | | | | ***AD*** | | | | ***RD*** | | | |
|---|---|---|---|---|---|---|---|---|---|---|---|---|---|---|---|---|
| | ***B (SE)*** | ***β*** | ***p*** | ***q*** | ***B (SE)*** | ***β*** | ***p*** | ***q*** | ***B (SE)*** | ***β*** | ***p*** | ***q*** | ***B (SE)*** | ***β*** | ***p*** | ***q*** |
| **Model 9: L CST** | | | | | | | | | | | | | | | | |
| | $R^2$ = .10** | | | | $R^2$ = .09** | | | | $R^2$ = .08** | | | | $R^2$ = .08** | | | |
| GA | .27 (.18) | .13 | .14 | .15 | .30 (.18) | .14 | .10 | .12 | .31 (.18) | .15 | .09 | .18 | .29 (.19) | .14 | .11 | .11 |
| Sex | .92 (.45) | .16 | .04 | .05 | .90 (.45) | .16 | .05 | .05 | .91 (.45) | .16 | .04 | .06 | .89 (.45) | .16 | .05 | .05 |
| PMA at scan | .02 (.19) | .01 | .92 | .92 | .12 (.20) | .06 | .55 | .85 | .16 (.19) | .08 | .41 | .76 | .10 (.20) | .05 | .62 | .80 |
| L CST | 9.55 (13.4) | .07 | .48 | .48 | 1.04 (2.85) | .03 | .72 | .72 | 2.89 (3.25) | .08 | .38 | .38 | .39 (2.65) | .01 | .89 | .88 |

*Note.* FA, fractional anisotropy; MD, mean diffusivity; AD, axial diffusivity; RD, radial diffusivity; GA, gestational age; PMA, postmenstrual age; SLF/AF, superior longitudinal fasciculus/arcuate fasciculus; IFOF, inferior fronto-occipital fasciculus; CST, corticospinal tract; R, right; L, left

*p<.05, **p<.01, ***p<.001

**Table S6.** No significant associations between neonatal mean and radial diffusivity and global language outcomes at age 2 years

| | *MD* | | | | *RD* | | | |
|---|---|---|---|---|---|---|---|---|
| | ***B (SE)*** | ***β*** | ***p*** | ***q*** | ***B (SE)*** | ***β*** | ***p*** | ***q*** |
| **Model 1: Corpus Callosum** | | $R^2 = .08^{**}$ | | | | $R^2 = .09^{**}$ | | |
| GA | 2.34 (1.05) | .18 | .03 | .04 | 2.29 (1.04) | .18 | .03 | .04 |
| Sex | 5.25 (2.62) | .16 | .05 | .05 | 5.44 (2.60) | .16 | .04 | .05 |
| PMA at scan | .40 (1.19) | .34 | .73 | .73 | -.09 (1.16) | -.01 | .94 | .94 |
| Corpus Callosum | -44.6 (47.20 | -.09 | .35 | .35 | -65.0 (34.7) | -.17 | .06 | .06 |
| **Model 2: R SLF/AF** | | $R^2 = .07^{*}$ | | | | $R^2 = .07^{*}$ | | |
| GA | 2.19 (1.07) | .17 | .04 | .04 | 2.20 (1.07) | .17 | .04 | .04 |
| Sex | 5.17 (2.63) | .15 | .05 | .05 | 5.18 (2.63) | .15 | .05 | .05 |
| PMA at scan | .72 (1.11) | .06 | .52 | .66 | .74 (1.11) | .06 | .50 | .66 |
| R SLF/AF | -5.61 (9.52) | -.05 | .56 | .56 | -4.76 (8.90) | -.05 | .59 | .59 |
| **Model 3: L SLF/AF** | | $R^2 = .05^{*}$ | | | | $R^2 = .05^{*}$ | | |
| GA | 2.25 (1.07) | .18 | .04 | .04 | 2.24 (1.07) | .18 | .04 | .04 |
| Sex | 5.27 (2.63) | .16 | .05 | .05 | 5.27 (2.63) | .16 | .0 | .05 |
| PMA at scan | .87 (1.14) | .07 | .45 | .66 | .95 (1.15) | .07 | .46 | .66 |
| L SLF/AF | -2.32 (10.7) | -.02 | .83 | .83 | -2.65 (9.86) | -.03 | .79 | .79 |
| **Model 4: R Uncinate** | | $R^2 = .07^{*}$ | | | | $R^2 = .07^{*}$ | | |
| GA | 2.37 (1.06) | .19 | .03 | .04 | 2.33 (1.06) | .18 | .03 | .04 |
| Sex | 5.14 (2.63) | .15 | .05 | .05 | 5.22 (2.62) | .15 | .05 | .05 |

**Table S6.** No significant associations between neonatal mean and radial diffusivity and global language outcomes at age 2 years

| | *MD* | | | | *RD* | | | |
|---|---|---|---|---|---|---|---|---|
| | ***B (SE)*** | ***β*** | ***p*** | ***q*** | ***B (SE)*** | ***β*** | ***p*** | ***q*** |
| PMA at scan | .59 (1.15) | .05 | .27 | .66 | .64 (1.16) | .05 | .58 | .66 |
| R Uncinate | -25.6 (34.1) | -.07 | .59 | .59 | -18.5 (30.3) | -.06 | 54 | .54 |
| **Model 5: L Uncinate** | $R^2$ = .07* | | | | $R^2$ = .17*** | | | |
| GA | 2.25 (1.06) | .18 | .04 | .04 | 2.29 (1.05) | .18 | .03 | .04 |
| Sex | 5.26 (2.62) | .16 | .05 | .05 | 5.28 (2.62) | .16 | .05 | .05 |
| PMA at scan | 1.25 (1.13) | .10 | .27 | .66 | 1.19 (1.13) | .10 | .30 | .66 |
| L Uncinate | 18.4 (33.9) | .05 | .59 | .59 | 12.1 (30.8) | .04 | .69 | .69 |
| **Model 6: R IFOF** | $R^2$ = .07* | | | | $R^2$ = .07* | | | |
| GA | 2.31 (1.05) | .18 | .03 | .04 | 2.32 (1.06) | .18 | .03 | .04 |
| Sex | 5.50 (2.62) | .16 | .04 | .05 | 5.39 (2.62) | .16 | .04 | .05 |
| PMA at scan | 1.76 (1.27) | .14 | .17 | .66 | 1.50 (1.28) | .12 | .24 | .66 |
| R IFOF | 32.0 (31.5) | .10 | .31 | .31 | 16.5 (25.3) | .07 | .52 | .52 |
| **Model 7: L IFOF** | $R^2$ .07* | | | | $R^2$ = .08** | | | |
| GA | 2.27 (1.05) | .18 | .03 | .04 | 2.30 (1.05) | .18 | .03 | .04 |
| Sex | 5.42 (2.62) | .16 | .04 | .05 | 5.29 (2.62) | .16 | .05 | .05 |
| PMA at scan | 1.88 (1.29) | .15 | .15 | .66 | 1.68 (1.29) | .14 | .20 | .66 |
| L IFOF | 37.7 (33.7) | .11 | .26 | .26 | 23.2 (26.9) | .09 | .39 | .39 |
| **Model 8: R CST** | $R^2$ = .08* | | | | $R^2$ = .07* | | | |

**Table S6.** No significant associations between neonatal mean and radial diffusivity and global language outcomes at age 2 years

| | *MD* | | | | *RD* | | | |
|---|---|---|---|---|---|---|---|---|
| | ***B (SE)*** | ***β*** | ***p*** | ***q*** | ***B (SE)*** | ***β*** | ***p*** | ***q*** |
| GA | 2.23 (1.07) | .18 | .04 | .04 | 2.19 (1.07) | .17 | .04 | .04 |
| Sex | 5.28 (2.62) | .16 | .05 | .05 | 5.28 (2.62) | .16 | .05 | .05 |
| PMA at scan | .83 (1.14) | .07 | .46 | .66 | .75 (1.14) | .06 | .51 | .66 |
| R CST | -4.98 (16.5) | -.03 | .76 | .76 | -7.22 (15.4) | -.04 | .64 | .64 |
| **Model 9: L CST** | $R^2 = .08$* | | | | $R^2 = .07$* | | | |
| GA | 2.25 (1.07) | .18 | .04 | .04 | 2.20 (1.07) | .17 | .04 | .04 |
| Sex | 5.29 (2.62) | .16 | .05 | .05 | 5.29 (2.62) | .16 | .05 | .05 |
| PMA at scan | .88 (1.16) | .07 | .45 | .66 | .76 (1.16) | .06 | .51 | .66 |
| L CST | -3.35 (16.8) | -.02 | .84 | .84 | -6.46 (15.6) | -.04 | .68 | .69 |

*Note.* MD, mean diffusivity; RD, radial diffusivity; GA, gestational age; PMA, postmenstrual age; SLF/AF, superior longitudinal fasciculus/arcuate fasciculus; IFOF, inferior fronto-occipital fasciculus; CST, corticospinal tract; R, right; L, left

*p<.05, **p<.01, ***p<.001

**Table S7.** Associations between corpus callosum axial diffusivity and language outcomes after accounting for prenatal Social Disadvantage

| Predictors | *Global Language* | | | *Receptive Language* | | | *Expressive Language* | | |
|---|---|---|---|---|---|---|---|---|---|
| | ***B (SE)*** | **$\beta$** | ***p*** | ***B (SE)*** | **$\beta$** | ***p*** | ***B (SE)*** | **$\beta$** | ***p*** |
| **Step 1** | **$R^2$ = .25 ***** | | | **$R^2$ = .27 ***** | | | **$R^2$ = .17 ***** | | |
| GA | 1.41 (.91) | .11 | .12 | .32 (.17) | .13 | .06 | .15 (.16) | .07 | .36 |
| Sex | 4.40 (2.36) | .13 | .06 | .78 (.45) | .12 | .09 | .77 (.42) | .14 | .07 |
| PSD | -7.61 (1.24) | -.44 | <.001 | -1.51 (.24) | -.45 | <.001 | -1.07 (.22) | -.37 | <.001 |
| **Step 2** | **$\Delta R^2$ = .01** | | | **$\Delta R^2$ = .01** | | | **$\Delta R^2$ = .01** | | |
| GA | 1.08 (.97) | .09 | .27 | .24 (.18) | .10 | .20 | .12 (.17) | .06 | .49 |
| Sex | 4.83 (2.39) | .14 | .05 | .86 (.45) | .13 | .06 | .83 (.42) | .15 | .04 |
| PSD | -7.22 (1.28) | -.42 | <.001 | -1.42 (.24) | -.43 | <.001 | -1.02 (.23) | -.35 | <.001 |
| PMA at scan | .83 (.96) | .07 | .39 | .23 (.18) | .10 | .22 | .06 (.17) | .03 | .72 |
| CC AD | 53.3 (44.6) | .09 | .23 | 10.2 (8.44) | .09 | .23 | 7.63 (7.88) | .08 | .33 |

*Note.* GA, gestational age; PSD, prenatal Social Disadvantage; PMA, postmenstrual age; CC AD, corpus callosum axial diffusivity

*p<.05, **p<.01, ***p<.001

**Table S8.** Mediation analysis of prenatal Social Disadvantage, neonatal corpus callosum fractional anisotropy and axial diffusivity, and global/receptive language outcomes at age 2 years

| **Path** | **Path description** | **B** | **SE (HC3)** | ***t*** | **95% CIs** |
|---|---|---|---|---|---|
| **Global Language** | | | | | |
| ***Corpus Callosum Fractional Anisotropy (CC FA)*** | | | | | |
| Total effect (c) | SD → Global Language | -7.61 | 1.24 | -6.12 | **-10.07 – -5.16** |
| Direct effect (c’) | SD → Global Language with CC FA in the model | -7.21 | 1.21 | -6.97 | **-9.60 – -4.83** |
| Indirect effect (ab) | SD → CC FA → Global Language | -0.34 | 0.28 | - | -0.96 – 0.15 |
| ***Corpus Callosum Axial Diffusivity (CC AD)*** | | | | | |
| Total effect (c) | SD → Global Language | -7.61 | 1.24 | -6.12 | **-10.07 – -5.16** |
| Direct effect (c’) | SD → Global Language with CC AD in the model | -7.22 | 1.22 | -5.91 | **-9.63 – -4.80** |
| Indirect effect (ab) | SD → CC AD → Global Language | -0.33 | 0.31 | - | -0.99 – 0.25 |
| **Receptive Language** | | | | | |
| ***Corpus Callosum Fractional Anisotropy (CC FA)*** | | | | | |
| Total effect (c) | SD → Receptive Language | -1.51 | 0.24 | -6.38 | **-1.97 – -1.04** |
| Direct effect (c’) | SD → Receptive Language with CC FA in the model | -1.42 | 0.24 | -6.04 | **-1.88 – -0.96** |
| Indirect effect (ab) | SD → CC FA → Receptive Language | -0.07 | 0.04 | - | -0.19 – 0.03 |
| ***Corpus Callosum Axial Diffusivity (CC AD)*** | | | | | |

**Table S8.** Mediation analysis of prenatal Social Disadvantage, neonatal corpus callosum fractional anisotropy and axial diffusivity, and global/receptive language outcomes at age 2 years

| **Path** | **Path description** | **B** | **SE (HC3)** | ***t*** | **95% CIs** |
|---|---|---|---|---|---|
| Total effect (c) | SD → Receptive Language | -1.51 | 0.24 | -6.38 | **-1.97 – -1.04** |
| Direct effect (c') | SD → Receptive Language with CC AD in the model | -1.42 | 0.24 | -5.86 | **-1.90 – -0.94** |
| Indirect effect (ab) | SD → CC AD → Receptive Language | -0.07 | 0.06 | - | -0.18 – 0.04 |

*Note.* B, unstandardized co-efficient; SE (HC3), standard error with HC3 heteroscedasticity-consistent inference; CIs, Confidence Intervals; CC, corpus callosum; FA, fractional anisotropy; AD, axial diffusivity.

Models adjusted for sex, gestational age at birth and postmenstrual age at MRI scan. 95% CIs for indirect effects calculated with bootstrapping (5000 samples).

**No Interaction between CC FA and PSD on Global or Receptive Language Outcomes by Age 2 Years**

Formal moderation analysis was used to test the interaction between PSD and neonatal CC FA on global and receptive language outcomes to examine whether associations between CC microstructure and language outcomes varied as a function of PSD. GA at birth, sex, and PMA at MRI scan were included as covariate factors. As shown in Table S9, there was no interaction between PSD and CC FA on either global or receptive language scores by age 2 years (p>.05). Null results were also obtained when we tested an interaction between neonatal CC FA and year-2 Social Disadvantage to explore whether infants with microstructural alterations may be more sensitive to postnatal disadvantage (p>.05, see Main Text).

**Table S9.** Formal moderation analyses with corpus callosum fractional anisotropy and prenatal Social Disadvantage

| | ***Global Language*** | | | ***Receptive Language*** | | |
|---|---|---|---|---|---|---|
| | ***B (SE)*** | ***t*** | ***p*** | ***B (SE)*** | ***t*** | ***p*** |
| | $R^2$ **= .29 ***** | | | $R^2$ **= .31 ***** | | |
| GA | 1.22 (.95) | 1.29 | .20 | .26 (.18) | 1.46 | .15 |
| Sex | 5.08 (2.34) | 2.17 | .03 | .91 (.44) | 2.05 | .04 |
| PSD | -6.91 (1.25) | -5.51 | **<.001** | -1.36 (.24) | -5.74 | **<.001** |
| PMA at scan | -.32 (.98) | -.33 | .75 | .00 (.19) | .03 | .98 |
| CC FA | 104.4 (45.0) | 2.32 | **.02** | 20.1 (8.52) | 2.36 | **.02** |
| PSD x CC FA | -62.5 (41.7) | -1.50 | .14 | -11.9 (7.89) | -1.51 | .13 |

*Note.* GA, gestational age; PSD, prenatal Social Disadvantage; PMA, postmenstrual age; CC FA, corpus callosum fractional anisotropy

*p<.05, **p<.01, ***p<.001

**Specificity of Corpus Callosum Microstructure Findings for Language Outcomes**

To investigate if findings regarding neonatal CC FA and CC AD were specific to language outcomes, regression models were repeated using Bayley-III Cognitive Composite scores as the dependent variable. As shown in Table S14, neonatal CC FA and AD were not related to Bayley-III Cognitive Composite scores (p≤.10).

**Table S10.** Associations between corpus callosum fractional anisotropy and axial diffusivity and cognitive outcome at age 2 years

| | ***Global Language*** | | | | | |
|---|---|---|---|---|---|---|
| | ***B (SE)*** | ***β*** | ***p*** | ***B (SE)*** | ***β*** | ***p*** |
| | *$R^2$ = .11*** | | | *$R^2$ = .11*** | | |
| GA | 1.96 (.77) | .21 | .01 | 1.83 | .20 | .02 |
| Sex | 3.37 (1.92) | .15 | .06 | 3.80 | .15 | .05 |
| PMA at scan | .46 (.81) | .05 | .57 | 1.35 | .15 | .08 |
| CC FA | 62.0 (36.8) | .14 | .09 | - | - | - |
| CC AD | - | - | - | 58.1 | .13 | .10 |

*Note.* GA, gestational age; PMA, postmenstrual age; CC FA, corpus callosum fractional anisotropy

*p<.05, **p<.01, ***p<.001

**Sensitivity Analyses After Excluding Children with Severe Cognitive Delay and After Excluding Children Born Preterm.**

To ensure that results were not driven by a small group of children either with severe cognitive delay (Bayley-III Cognition < 70, *n* = 17, 11%) or born preterm (<37 weeks GA, *n* = 15, 9%), key regression models were re-run excluding these participants. As shown in Table S15 below, main study findings were unchanged, such that PSD and CC microstructure were related with language outcomes.

**Table S11.** Summary of regression analyses excluding children with severe cognitive delay or born preterm

| | ***Global Language*** | | | ***Receptive Language*** | | | ***Expressive Language*** | | |
|---|---|---|---|---|---|---|---|---|---|
| | ***B (SE)*** | ***β*** | ***p*** | ***B (SE)*** | ***β*** | ***p*** | ***B (SE)*** | ***β*** | ***p*** |
| **Excluding children with severe cognitive delay** | | | | | | | | | |
| GA | .62 (.94) | -.05 | .51 | .18 (.18) | .07 | .32 | .02 (.17) | .01 | .91 |
| Sex | 5.57 (2.29) | .17 | .02 | .98 (.43) | .16 | .03 | .87 (.42) | .18 | .02 |
| PSD | -7.75 (1.16) | -.48 | **<.001** | -1.54 (.22) | -.50 | **<.001** | -1.08 (.21) | -.39 | **<.001** |
| PMA at MRI | -1.24 (.96) | -.10 | .20 | -.16 (.18) | -.07 | .38 | -.25 (.18) | -.12 | .16 |
| CC FA | 133.4 (43.2) | .24 | **.01** | 25.7 (8.17) | .24 | **.01** | 19.6 (7.90) | .21 | **.02** |
| **Excluding children born preterm** | | | | | | | | | |
| GA | -.80 (1.30) | -.05 | .54 | -.10 (.25) | -.03 | .70 | -.19 (.23) | -.07 | .41 |
| Sex | 5.57 (2.40) | .17 | .02 | 1.00 (.46) | .15 | .03 | .94 (.43) | .17 | .03 |
| PSD | -8.16 (1.27) | -.48 | **<.001** | -1.59 (.24) | -.49 | **<.001** | -1.17 (.23) | -.41 | **<.001** |
| PMA at MRI | .10 (1.02) | .01 | .92 | .07 (.19) | .03 | .72 | -.02 (.18) | -.01 | .89 |

**Table S11.** Summary of regression analyses excluding children with severe cognitive delay or born preterm

| | ***Global Language*** | | | ***Receptive Language*** | | | ***Expressive Language*** | | |
|---|---|---|---|---|---|---|---|---|---|
| | ***B (SE)*** | ***β*** | ***p*** | ***B (SE)*** | ***β*** | ***p*** | ***B (SE)*** | ***β*** | ***p*** |
| CC FA | 106.1 (45.6) | .19 | **.02** | 20.1 (8.68) | .18 | **.02** | 15.7 (8.17) | .16 | .06 |

*Note.* CC FA, corpus callosum fractional anisotropy; PSD, prenatal Social Disadvantage
